# Virtual Epileptic Patient (VEP): Data-driven probabilistic personalized brain modeling in drug-resistant epilepsy

**DOI:** 10.1101/2022.01.19.22269404

**Authors:** Huifang E Wang, Marmaduke Woodman, Paul Triebkorn, Jean-Didier Lemarechal, Jayant Jha, Borana Dollomaja, Anirudh Nihalani Vattikonda, Viktor Sip, Samuel Medina Villalon, Meysam Hashemi, Maxime Guye, Julia Scholly, Fabrice Bartolomei, Viktor Jirsa

## Abstract

One-third of 50 million epilepsy patients worldwide suffer from drug resistant epilepsy and are candidates for surgery. Precise estimates of the epileptogenic zone networks (EZNs) are crucial for planning intervention strategies. Here, we present the Virtual Epileptic Patient (VEP), a multimodal probabilistic modeling framework for personalized end-to-end analysis of brain imaging data of drug resistant epilepsy patients. The VEP uses data-driven, personalized virtual brain models derived from patient-specific anatomical (such as T1-MRI, DW-MRI, and CT scan) and functional data (such as stereo-EEG). It employs Markov Chain Monte Carlo (MCMC) and optimization methods from Bayesian inference to estimate a patient’s EZN while considering robustness, convergence, sensor sensitivity, and identifiability diagnostics. We describe both high-resolution neural field simulations and a low-resolution neural mass model inversion. The VEP workflow was evaluated retrospectively with 53 epilepsy patients and is now being used in an ongoing clinical trial (EPINOV).

## Main

Epilepsy is among the most common neurological disorders. The identification of the brain regions (epileptogenic zone, EZ) involved in the genesis of seizures is a necessary step before any epilepsy surgery. This identification is based on non-invasive data (MRI, PET, EEG, MEG) and, frequently, on the basis of intracerebral recordings, in particular stereoelectroencephalography (SEEG). Numerous works have shown that epileptogenic regions are more often organized in a network than originating from a single epileptic focus^1,2^. SEEG data indicate that epileptogenic networks follow a hierarchical organization with maximal epileptogenicity in the epileptogenic zone network (EZN)^1^. Many methods have been proposed to quantify these EZNs. The epileptogenicity index (EI) quantifies the EZN based on both the spectral and temporal properties of SEEG signals^3^. Subsequently, connectivity EI (cEI)^4^ combines connectivity features with EI for better prediction, especially of seizures with slow onset patterns. A statistical parametric map method was proposed to map EI from SEEG signals anatomically onto the patient’s MRI^5^. Three biomarkers introduced for EZ identification include fast activity in the frequency band of 80-120 Hz together with a very slow transient polarizing shift and voltage depression^6^. A support vector machine was used to identify a specific time-frequency pattern in the EZ that included a combination of sharp transients or spikes with preceding multiband fast activity concurrent with suppression of lower frequencies^7^. Clinical practice requires sophisticated diagnostic approaches and the expertise of trained clinicians to integrate and analyze information from multiple modalities and then determine the EZN. Therefore, methods of causal inference are needed to provide better EZN estimates by naturally integrating multiple modalities to identify the underlying latent mechanisms, which can provide better estimates than those obtained by observation alone. Virtual brains are data-driven dynamical brain network models that include in their composition non-invasively obtained connectivity data of individual subjects. The Virtual Brain (TVB)^8^ is a neuroinformatics platform for the construction and simulation of virtual brains that can provide the required functionality for EZN estimation and make it possible to perform causal inference on the full brain scale.

Here, we introduce the VEP workflow, which uses personalized virtual brain models and machine learning methods by integrating epilepsy patient-specific anatomical with functional data to aid in clinical decision-making by estimating the EZN and further optimizing the surgical strategy. The full brain models are built in a patient-specific space defined by T1-weighted magnetic resonance imaging data (T1-MRI). Neural models are built for each node, which is each vertex out of 260,000 in high-resolution neural field models (NFM) and one brain region out of 162 in the low-resolution neural mass models (NMM). Diffusion-weighted MRI (DW-MRI) is used to define a patient’s specific connectivity between the nodes. We previously demonstrated that patient-specific connectivity information can improve the estimation of parameters^9,10^. Inferring virtual brain model parameters such that the simulated data match the patient’s seizure organization further increases the patient specificity. This inference process identifies the EZN using techniques from machine learning, in particular through Markov chain Monte Carlo (MCMC) sampling. This method generates many random samples of model parameters and virtual brain simulations and evaluates these for consistency with empirical data (i.e., the patient’s functional data SEEG recording). Inferences can be further improved by integrating prior knowledge about the spatial distribution of the EZN, such as that obtained from the spatial extent of lesions in MRI data or seizure semiology, to reduce uncertainty and to bias the non-identifiable parameter estimation. After the first proof-of-concept of the VEP^11^, we studied and tested different modeling methods^12–15^, model inversion methods^16–19^, and surgery strategies^20,21^. Here, we present a complete and extensively tested VEP workflow that can be used in clinical practice. This also provides a foundation for further improvements in both brain modeling and epilepsy studies.

## Results

### VEP workflow

VEP assists in the identification of the EZN based on a personalized whole brain model. The model defines the brain as a network of regions, each representing a node in the brain network (see Fig. 1 for the workflow and Supplementary Fig. 1 for the flowchart.). These regions are delineated by the VEP atlas, a cortical and sub-cortical parcellation of the brain developed specifically for use in epileptology and functional neurosurgery^22^. The VEP atlas considers region sizes adapted to EZN diagnostics for improved performance of model inversion techniques. Using geometric information and neuroanatomical conventions, brain regions can be automatically labeled, based on the patients’ T1 weighted MR images (T1-MRI). Connections between regions are estimated through streamline tractography from diffusion-weighted magnetic resonance imaging (DW-MRI). Together with the brain parcellation from the T1-MRI, this method can provide a patient-specific structural connectivity matrix, which defines the links between the nodes of the network. The dynamics of each node are defined by a NMM, a system of nonlinear differential equations that represent the neural dynamics in that brain region. In this work, we used the Epileptor^23^, a phenomenological NMM, capable of simulating seizure-like activity.

**Fig. 1.**
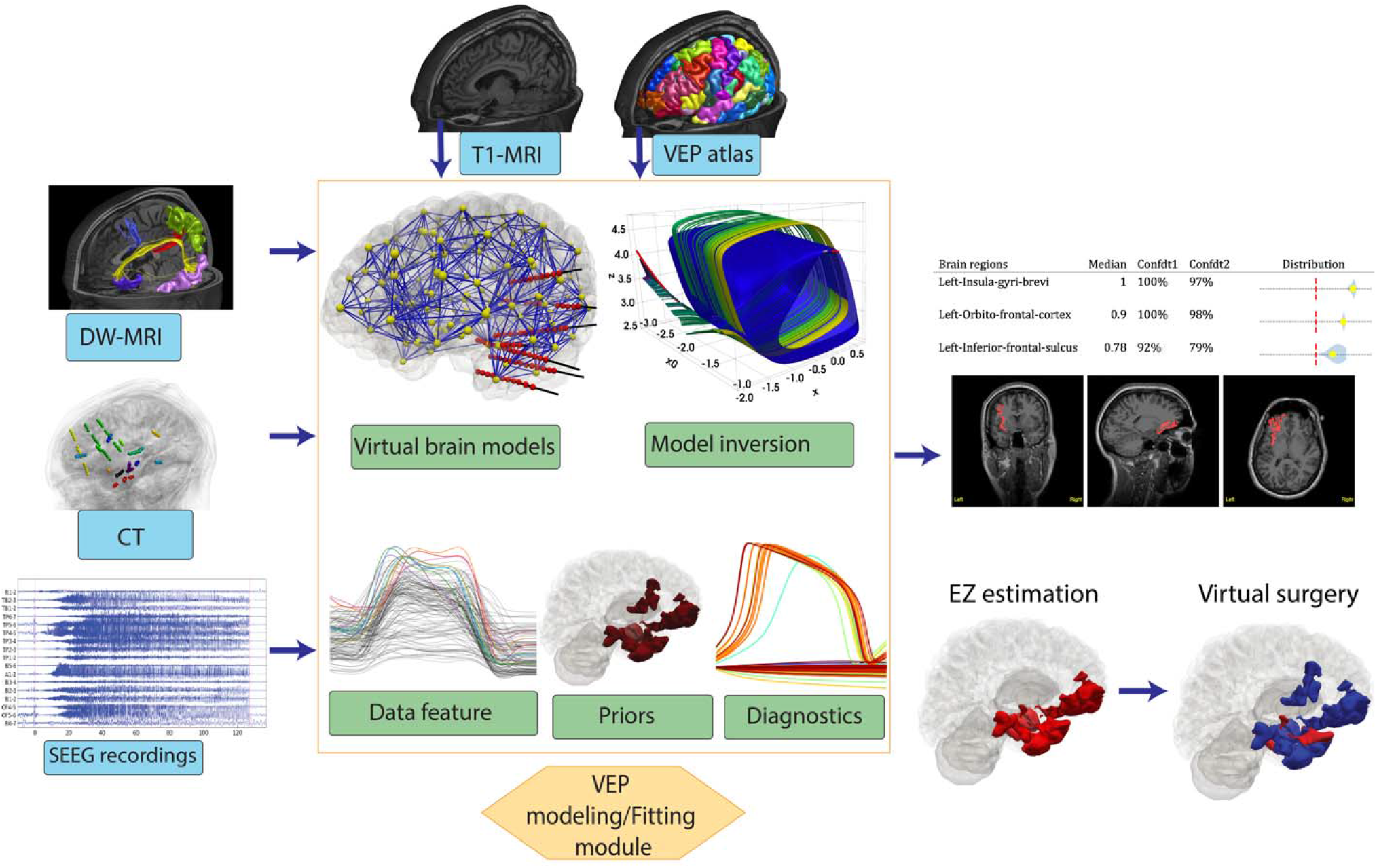
Workflow of the Virtual Epileptic Patient (VEP). A T1 weighted MRI (T1-MRI) is used to extract the individual brain geometry and to define distinct brain regions according to the VEP atlas. Tractography is performed on diffusion weighted MRI (DW-MRI) data to estimate the length and density of the white matter fibers. These are grouped according to regions obtained from the atlas to derive a structural connectivity matrix that specifies the connection strength between brain regions. A post-SEEG-implantation CT scan is used to find the exact locations of the SEEG electrodes and construct the source-to-sensor map using the so-called gain matrix. Neural mass models at each source location are connected through the connectivity matrix, and the neural source activity are simulated. The gain matrix maps the simulated source activity to the corresponding SEEG signals. Bayesian inference methods are used to estimate the patient-specific parameters of the model based on the data features extracted from SEEG signals and priors from added knowledge, such as analysis of the SEEG data or clinical hypotheses. The output is the suggested EZ network, which can be communicated by a distribution table and heatmap in the T1-MRI data.

The signals generated by Epileptor models on the brain region level are called source signals. The measured signals from the SEEG electrodes are called sensor signals. To map the simulated sources from the brain regions to the SEEG sensors, the electromagnetic forward problem needed to be solved. Localizing the implanted SEEG electrodes in patient-specific space uses the co-registration of post-SEEG-implant computed tomography (CT) with the T1-MRI image. The source-to-sensor map, the so-called gain matrix, is a function of the distance between the sources and sensors^24^. Brain regions, the connectivity matrix, and the gain matrix are all defined in the patient-specific brain space, which are determined by the T1-MRI images. Note that, in the VEP pipeline the source space covers the whole brain, specifically, 162 brain regions defined by the VEP atlas in the NMM, whereas the sensors only sample part of the brain, depending on the patient-specific SEEG implantation scheme.

The model inversion module allows us to infer the unknown model parameters by fitting the SEEG data features. The VEP workflow includes two independent model inversion methods based on Bayesian inference, namely the optimization and sampling pipelines, which complement each other. The optimization pipeline uses the LBFGS algorithm^25^, given prior information and the likelihood function of the data, to obtain a (local) maximum of the posterior distribution estimate of the model parameters, so-called a maximum a posteriori (MAP). The low computational costs and high robustness of this MAP method allow us to measure the sensitivity of the estimated results in terms of the sampled SEEG sensor space. To do this, 100 MAP estimates are obtained on datasets with a random sensor removed, resulting in a distribution of epileptogenicity values (EVs) for each region. In the sampling pipeline, full Bayesian inference uses a self-tuning variant of the Hamiltonian Monte Carlo (HMC) algorithm to estimate the potential multimodal posterior distributions of the parameters of the personalized NMMs^26,27^. Multiple chains start from different random initial conditions to sample the model parameters from their respective posterior distributions. While HMC handles large parameter space well, its implementations still struggle with the high-dimensional and unfavorable posterior geometries, so a reparameterization of the regional parameters is used to facilitate an efficient exploration of the posterior distribution over the reparameterized configuration space in terms of computational time and convergence diagnostics.

The inputs of the model inversion module are the data feature and a prior. The envelope of signal power (high-pass filtered signal followed by envelope smoothing) from empirical SEEG recordings serves as the data feature. We illustrate the data features from the two patients with the most common seizure onset patterns in Fig. 2**a**,**b** as well as three patients with other patterns in Supplementary Fig. 2. The prior can be estimated or defined by added knowledge, such as the MRI-visible lesions or the clinical EZ hypothesis based on a combination of clinical evidences or extended data analysis from the SEEG signal. We also designed prior estimate algorithms based on delay information from filtered SEEG signals in multiple frequency bands during seizure onset. The VEP workflow can use the different priors to provide a complete picture as the outcome from the model-based inference.

**Fig. 2.**
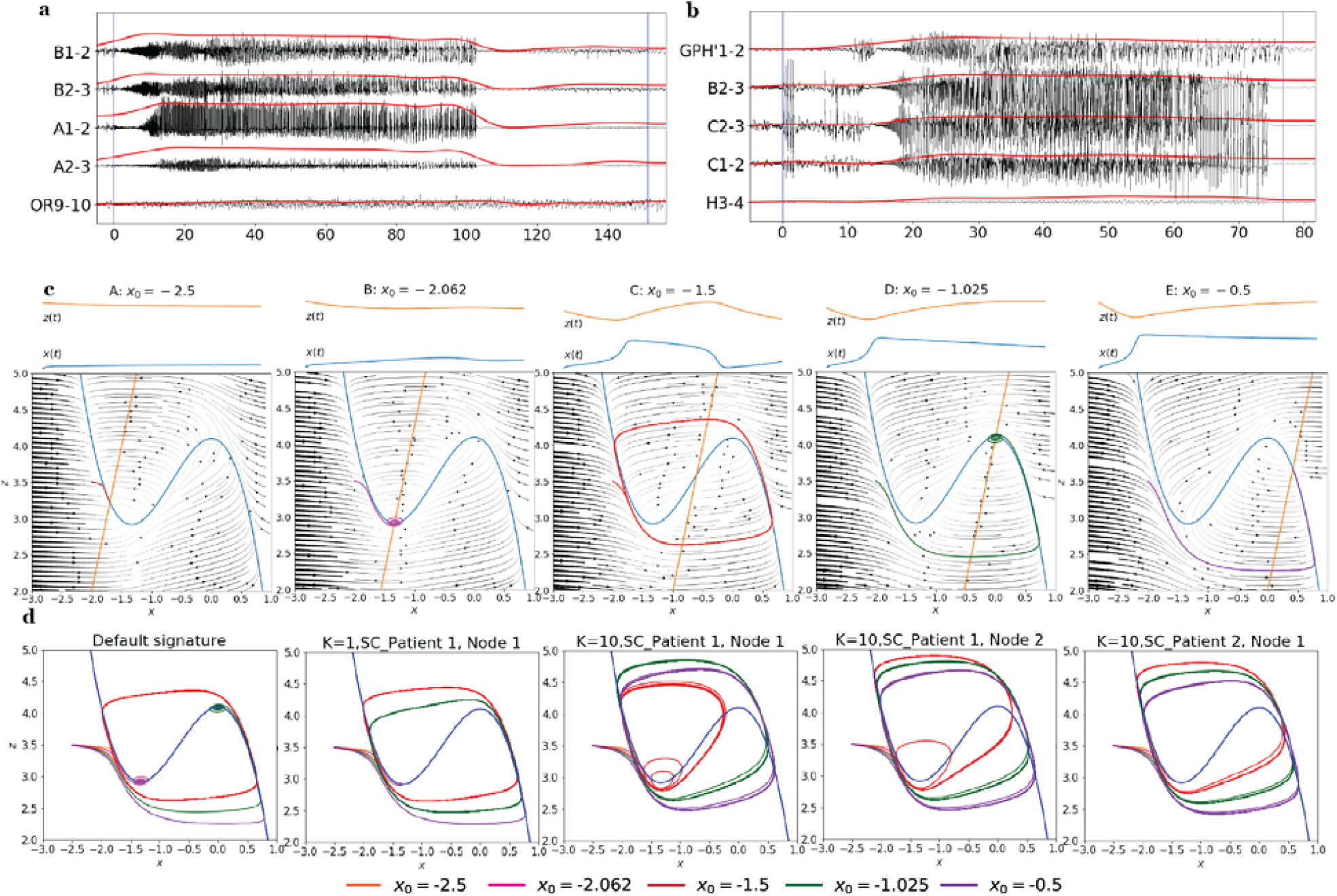
Data feature extraction and bifurcation analysis of the Epileptor. **a**,**b**, Raw SEEG recordings (in black) were high-pass filtered to extract the high frequency components of the signal. A smoothed envelope function (in red) was calculated from the filtered signal, which served as the data feature for model inversion. The data feature precisely captured the onset and offset of the seizure. **a**, Low-voltage fast activity (LVFA), a seizure onset pattern, was found in 45.6% of patients^30^. **b**, Slow wave prior to LVFA, a seizure onset pattern, was found in 15.5% of patients. **c**, The dynamics of the 2D Epileptor are shown in a phase plane. Graphs from left to right show the effect of an increase in the excitability parameter x_0_ on the dynamics. For values of x_0_<-2.062 the system settles into a stable fixed point. For values -2.062<x_0_<-1.025 the model generates a stable oscillation (see z(t) and x(t)), equivalent to the seizure time course of the data feature in **a**,**b**. For values −1.025 < x_0_ the Epileptor settles into a stable fixed point again. **d**, Trajectories of a single node in phase planes with 5 different x_0_ values under different network schemes. The dynamics not only depend on the value of x_0_, as depicted by the different colored trajectories but also on the connectivity scaling factor K, the specific node observed, and the patient specific connectome. Only a metric that takes into account all of these mentioned factors can make a meaningful prediction of the EZ.

The output of the model inversion module results in a distribution of EVs, which is determined by the key parameters from the model inversion and the patient’s structural connectivity. We will explain this important definition in more detail in the next subsection. An overview of the most relevant brain regions, according to the model estimate based on the EVs, is presented in the VEP clinical report. The report table contains the information from highlighted brain regions with the distribution of the estimated EVs, their median, and the confidence values. Additionally, the regions identified by the model as belonging to the EZN are represented in red in the three classical planes in the patient’s 3D T1-MRI images (called a heatmap), provided in a user-friendly viewer. Furthermore, the VEP workflow provides the therapeutic solution by a virtual surgical strategy based on the estimated EZN. Different surgical strategies can be tested to find a minimal resection area to achieve seizure-freedom for the patient while minimizing the risk of functional deficits.

### Definition of EZN from a modeling perspective

The concept of the EZ has developed along with advances in surgical experience and diagnostic techniques. There are two established and most frequently used definitions of the EZ. First, the EZ is defined as the connected brain regions involved in the primary organization of seizures^1,28^. Second, from a surgical perspective, the EZ is defined as the brain regions that are necessary and sufficient for initiating seizures and whose removal (or disconnection) guarantees the complete abolition of seizures^29^. The EZs are more often organized into a network termed the EZN^1,2^.

We introduce the definition of the EZN from the perspective of whole brain modeling. The Epileptor captures the dynamics of a seizure by comparing their simulated time series (top of Fig. 2**c**) with data features extracted from empirical data (Fig. 2**a**,**b** and Supplementary Fig. 2). Changing the excitability parameter of this model can cause a brain region to transition from a stable regime with trajectories trending towards a stable fixed point into an oscillatory or seizing regime with a limit cycle manifold; see Fig. 2**c** for an illustration of what happens in the absence of connectivity. In brain network models, the specific connectivity plays an important role in the dynamics of the system. At each node in the brain network, the bifurcation scheme and its corresponding limit-cycle manifold vary depending on the global connectivity between brain regions (Fig. 2**c-d**). We varied the excitability parameter of a given node under different connectivities when all the other nodes were in the healthy regions. We considered the three factors that impact the connection strength between nodes: differences in the connectome between patients, the connectivity scaling parameter *K*, and connections of the node of interest within the network. Fig. 2**d** demonstrates that these three connectivity factors all influence the seizure dynamics of a single node. Thus, the definition of the EZN on which we based our mathematical definition in our modeling system is those brain regions with a high degree of excitability under a given connectivity configuration that can lead to a seizure. We calculated the EV for each region based on the onset delays of the optimized or sampled model of seizure activity at the source level (see online Method). This considers both the degree of excitability and the specific connectivity.

### Validation on high-resolution synthetic data

Synthetic data can provide the ground-truth of the EZN, which we cannot obtain from empirical data. Thus, synthetic data is a useful tool for validating VEP pipelines by allowing investigators to compare the parameters of models used for the simulations with those inferred from the model inversion module. The simulation module of the VEP workflow for generating the synthetic testing data includes both NMMs in low space resolution and NFMs in high space resolution, where each node element of the model represents an average of ∼16 cm^2^ and ∼1 mm^2^ of the cortical surface, respectively. NMMs have proven their efficiency in capturing the main features of brain functional behaviors by accounting for interactions between brain regions^11^. In epilepsy, they can capture the main features of seizure generation and propagation while considering the underlying connectivity of the network characteristics of seizures. Their low computational cost makes model inversion and parameter inference possible. NMMs can be used for both the simulation and model inversion modules.

NFMs implement field equations representing neural activity in continuous space^31^, increasing the resolution by a factor of 1000 compared with the discrete point representation of brain regions in neural mass models. Neural fields allow the use of detailed connectivity, considering both local, between neighboring points on the field, and global connections, along white matter fibers. Another important advantage of neural field modeling is to provide more realistic source to sensor mapping by considering both the orientation and distance between the dipoles and the sensor. Supportive evidence comes from the experimental literature, which has shown that the current dipole is mainly attributable to pyramidal cells in the cortical grey matter and is aligned perpendicular to the surface^32^.

We validated the VEP using synthetic data generated by the NFMs (see Fig. 3). We used 6D Epileptor on each of the 260,000 vertices representing the cortical mesh. The EZN was set to cover 3 brain regions in the left hemisphere. Fig. **3a** illustrates the time series in the selected source signals and the signal power, which is mapped based on the brain high-resolution meshes. We observed complex spatial patterns within a brain region in which the seizure onset and offset times were not necessarily synchronized. We generated synthetic SEEG sensor data using the gain matrix while considering both the orientation and distance of the dipoles with respect to the sensors (Fig. 3**b**). The envelope function of the synthetic SEEG data provided the data feature for the VEP model inversion. Here the VEP inversion module was based on the NMM, with a lower dimensional model (2D Epileptor), which captures the envelope function, and a low-resolution gain matrix that considers only the distance between the nodes and sensors (Fig. 3**c**).

**Fig. 3.**
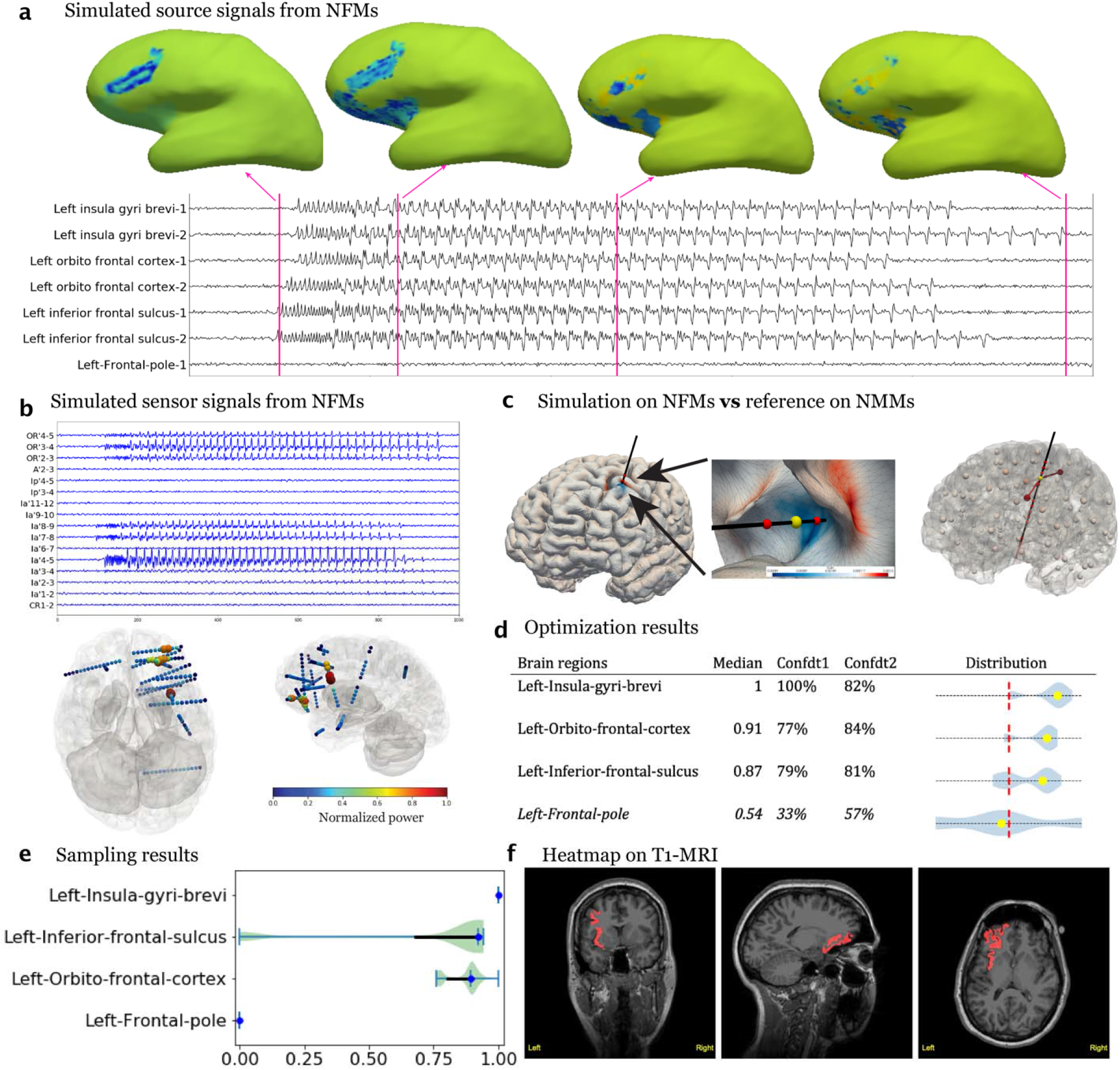
VEP validation using high-resolution synthetic data. **a**, Seizure dynamics were simulated using a NFM with an EZN of three given regions in the left hemisphere: left insula gyri brevi (L-IGB), left orbitofrontal cortex (L-OFC), and left inferior frontal sulcus (L-IFS). Neural activity is shown on the cortical mesh at 4 different time points. The time series of neural activity at 7 locations on the mesh are shown below the brain models in **a. b**, Synthetic SEEG data was derived by projecting the source activity using the gain matrix. Signal power at each SEEG contact, which is color-coded in a 3D cortical mesh, revealed strong activations in the frontal lobe. **c**, A NFM, which represents neural activity continuously in space and takes into account the electrical dipole orientation normal to the surface for the forward solution, was used for the simulation of the SEEG data. A computationally feasible reduced NMM was used for model inversion. **d**, A clinical report table shows the inferred EV distribution and obtained metrics from the optimization pipeline. The given threshold (0.5) is shown <u>by</u> the red dashed lines. **e**, The posterior distribution of EV values from the sampling pipeline. **f**, Identified EZN is shown in the patient-specific T1-MRI called the heatmap.

Based on the distribution of EVs, the EZN was declared. Both the optimization and sampling pipelines identified the ground-truth EZN, as shown in Fig. 3**d** and Fig. 3**e**, respectively. The optimization pipeline used the informative prior from our designed VEP prior algorithm. The clinical table based on the distribution of EVs shows the optimized results from datasets with different SEEG spatial samplings (Fig. 3**d**). The sampling pipeline used a noninformative prior in which all the brain regions followed the same prior distribution. The posterior distribution of EVs from the sampling pipeline was based on 16 chains starting from 8 different optimized initial conditions. Each HMC chain created 500 samples (Fig. 3**e**). The heatmap projected on the patient’s T1-MRI shows the spatial mapping of the EZN (sagittal, axial and coronal view images shown in Fig. 3**f** and an interactive html file in the Supplementary Materials).

### EZN estimation using empirical patient data

Next, we applied the VEP workflow to empirical data from a right-handed female patient, initially diagnosed with left frontal epilepsy. We first extracted the structural connectivity matrix (Supplementary Fig. 3) and source-to-sensor mapping (Supplementary Fig. 4) from individual T1-MRI, DW-MRI, and post-SEEG-implantation CT imaging data. Using these with the data features (Supplementary Fig. 5**a**) of SEEG seizure recordings as input, we ran both optimization and sampling pipelines. Fig. 4**a** shows the SEEG recording of one seizure and Fig. 4**b** demonstrates the distribution of the signal power among all the electrodes in a cortical mesh, with high activity in the frontal and insular cortex. A VEP prior algorithm calculated the sensor prior vector based on 52 different frequency bands from 10-110 Hz by taking into account the onset delay of the seizure in each channel (Supplementary Fig. 5**b**). Two different ways were used to project the prior value from the sensor to the source level: VEP-M directly maps the prior value of the sensor with the shortest distance to a given source (Supplementary Fig. 6); and VEP-W maps a weighted sum of the prior values of all sensors to each source, based on their distance (Supplementary Fig. 7) (see Methods). We ran the optimization pipeline with 4 different prior networks: VEP-M (Fig. 4**c**) and VEP-W (Supplementary Fig. 8**a**), clinical hypothesis (Supplementary Fig. 8**b**), and uninformative prior where all regions have the same probability distribution as the healthy regions (Supplementary Fig. 8**c**). The clinical hypothesis was obtained from neurologists (JS and FB) based on the EI index and other SEEG data, such as direct electrical stimulations. Summarizing the results from 4 runs, we argue that the left central operculum should be ruled out due to the prior networks. To investigate the influence of structural connectivity, we ran the optimization pipeline without considering any connection between brain regions, by taking into account an uninformative prior. In this example, we showed that both the L-IGB and L-OFC are identified to be part of the EZN only when this patient’s structural connectivity is considered (Supplementary Fig. 8**d**).

**Fig. 4:**
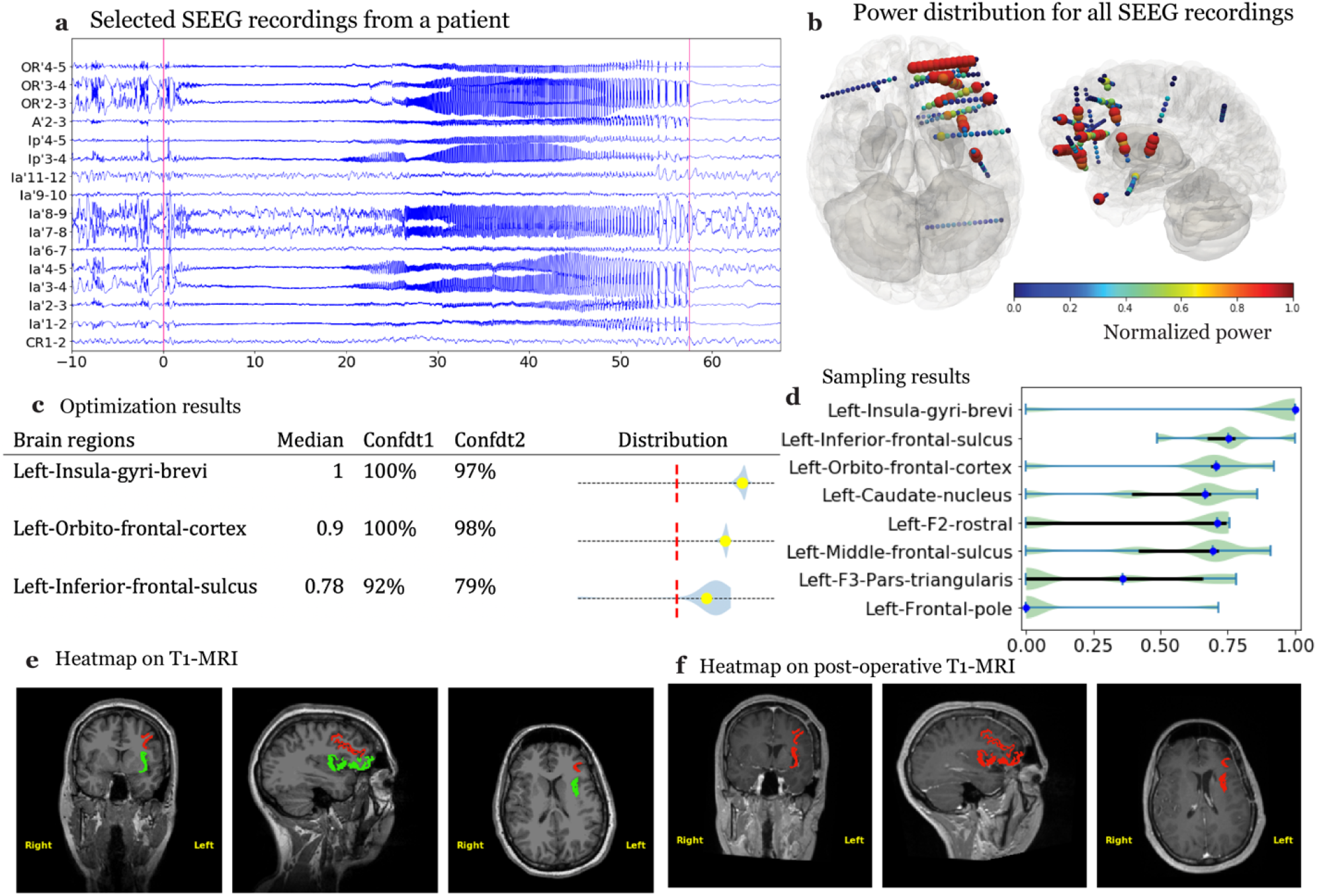
VEP results for clinical use case. **a**, Selected SEEG recordings from one seizure of a female patient. **b**, The distribution of the signal power across contacts inside the cortical mesh from two different perspectives. The sizes and the colors of the electrodes indicate signal power. Large size and red color indicate high power. **c**. Clinical report table from the optimization pipeline. **d**. Posterior distribution of the epileptogenicity value (higher value indicates higher probability for seizure) for 8 selected regions obtained from the sampling pipeline. **e**. Heatmap of the VEP results (in red) and clinical hypothesis (CH, in yellow), with overlapping regions in green, shown in a pre-operative T1-MRI. **f**. Heatmap of the VEP results (in red) shown in a post-operative T1-MRI.

In the sampling pipeline, we ran the HMC algorithms in 16 chains starting from 8 optimized initial conditions. Based on the sampled parameters of the NMM, we obtained the simulation that best matched the given data features, as measured by likelihood. We used the informative prior to bias the three brain regions identified as EZN by the optimization pipeline. The posterior distribution of 8 brain regions (Fig. 4**d)** confirmed the 3 brain regions identified by the optimization pipeline. Another four brain regions, seizing after these first 3 brain regions in the EZN, showed multimodal posterior probability distributions. The multimodality comes from the non-identifiability of the observable date feature, i.e., different parameter sets can lead to different patterns of latent source activity but still have a similar observable SEEG data in the sensor space. We analyzed four seizures of this patient and integrated the results of the four seizures to obtain the three brain regions as EZNs (Supplementary Fig. 9, and Methods). The EZN from the clinical hypothesis (L-IGB and L-OFC) and VEP atlas (L-IGB, L-OFC, and L-IFS) were overlaid onto the pre-surgical T1-MRI (Fig. 4**e**) and the post-operative T1-MRI (Fig. 4**f**). Two out of three of the VEP identified brain regions were also identified by the clinicians. The patient underwent resective surgery, which resulted in a reduction in seizure frequency but did not produce seizure freedom. Virtual surgery as an immediate application after the EZN estimation continues along this story line in the next subsection.

### VEP virtual surgery module

The estimation of the EZN provides a diagnostic result, whereas a therapeutic solution proposes the surgical intervention, such as the minimum number of brain regions that can be treated to allow seizure control while offering the best functional outcome. Furthermore, the VEP generative function can offer further advice about the type of intervention, such as stimulation, lesionectomy, or laser interstitial thermal therapy, by simulating the optimized brain models under different intervention conditions by varying its corresponding parameters. We refer to the simulation of different intervention hypotheses as “virtual surgery’’. Here we used the same patient as in the section before and worked with the inferred parameters to implement the identified three EZN regions in a VEP simulation. The simulated time-series and corresponding power distribution are shown in Fig. 5**a**. For this patient, we tested two virtual surgery hypotheses: removing the two brain regions identified in the clinician hypothesis (Fig. 5**b**) and removing six brain regions, as was done in the real surgery (Fig. 5**c**). Comparing the time-series before and after the virtual surgery (Fig. 5**b**,**c** vs Fig. 5**a**), the seizure reduction was obvious, but the patient was not seizure free, which is consistent with the real surgery outcome. Indeed, this patient was not seizure free after a surgery that removed the L-OFC and the L-IGB. Comparing the time-series between the two surgical hypotheses (Fig. 5**b** vs. Fig. 5**c**), we saw no notable difference between removing the 2 brain regions identified by the clinicians and removing 5 (2+3 additional) brain regions, as in the real surgery. The three additional brain regions in the real surgery were removed to provide access to the brain regions proposed by the neurologists.

**Fig. 5.**
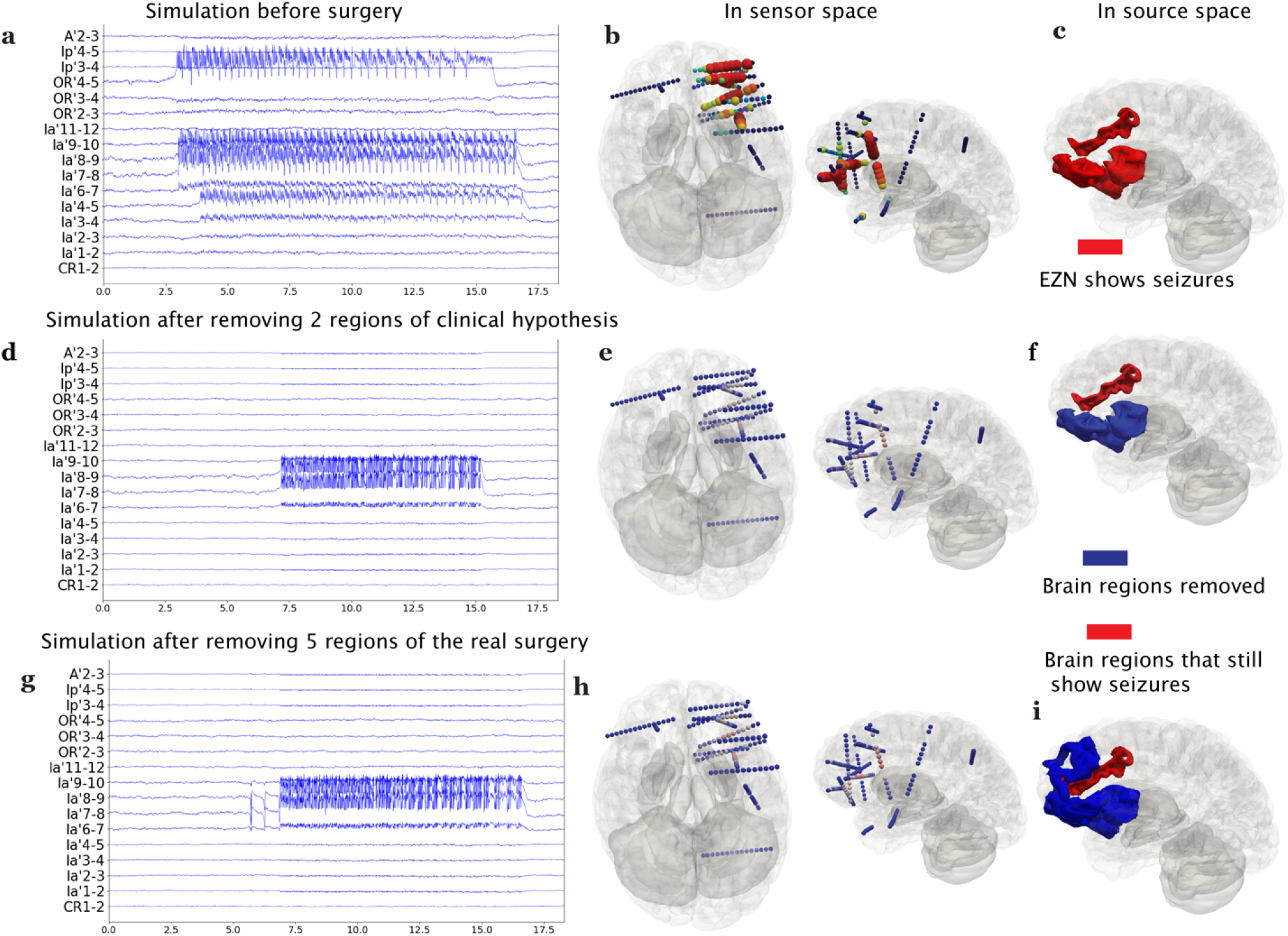
VEP virtual surgery example. **a**, Selected simulated SEEG time-series using the 6D-Epiletor with inferred parameters from the VEP results. **b**, Distribution of the signal power across contacts in a 3D brain from two different perspectives of the simulated data, as in **a. c**, EZN (highlighted in red) shows seizures in 3D brain. The EZN includes the left insula gyri brevi (L-IGB), left orbitofrontal cortex (L-OC), and left inferior frontal sulcus (L-IFS). **d, g** Selected simulated SEEG time-series after virtual surgery removing the two brain regions of the clinical hypothesis (**f**) and after removing the 5 regions of the real surgery (**i**). **e**,**h** Distribution of the signal power across SEEG contacts of virtual surgery simulation in 3D brain, corresponding to **c, f** respectively. **f**, 3D brain in the source space; the two regions (L-IGB and L-OC) in blue were virtually resected, and the red one (L-IFS) allowed the seizure activity to remain. **i**, 3D brain in the source space; the five regions (L-IGB, L-OC, left gyrus rectus, left F2 rostral, and left F3 pars orbitalis) in blue were resected both virtually and really, and the red one (L-IFS) allowed the seizure activity to remain.

Next, we investigated five virtual surgery hypotheses for another patient who had a resective surgery outcome of seizure freedom. We first built the simulated model using the VEP results for this patient with the three brain regions identified as the EZN. The simulated data is shown in Supplementary Fig. 10 **a**,**b**. The clinical hypothesis identified 7 regions in the right frontal and temporal regions as the EZN. If all of them would be removed, the virtual surgery suggested that there would be reduced seizure amplitude but still no seizure freedom (Supplementary Fig. 10 **c)**. Nineteen brain regions were removed in the real surgery, and the virtual surgery confirmed actual seizure freedom (Supplementary Fig. 10 **d)**. This is not surprising given such a large and redundant number of regions in the surgical resection. Then we considered two scenarios in terms of the global connectivity scale K. In the first scenario when the K was large, which could mean that the patient would be in a situation with fewer seizure triggers or with some anti-epilepsy drug, removing four regions was sufficient for seizure freedom (Supplementary Fig. 11 **a)**. In the second scenario in which K was smaller, removing a minimum of six regions led to seizure freedom (Supplementary Fig. 11**b**,**c)**, but removing 4 regions was not sufficient.

### VEP evaluation module

The performance of the VEP workflow in estimating the EZN for individual patients was quantified by validating the inferred EZN against the clinical hypothesis or postoperative MRI^3^ (see Fig. 6**a**). The commonly used seven metrics for the validation are based on true/false - positive/negative EZ classifications for each brain region based on presurgical evaluation (Fig. 6**b**). Compared with the clinical hypothesis, the predictive power of VEPs for a cohort of 53 retrospective patients (Fig. 6**b**) showed good precision (mean: 0.613) and acceptable recall (mean: 0.478) when a fixed threshold (0.5) was set for all patients. We also calculated the F_0.5_ (mean: 0.573), which is the weighted mean of the precision and recall with double weight on precision. If we used a personalized threshold, we obtained a higher precision (mean: 0.769) and F_0.5_ (mean: 0.652). Also, we introduced two threshold-free measures: the average precision score (APS: with mean 0.443) and the area under ROC curve (AUC with mean 0.761). Although a discrepancy existed between the VEP and the clinical hypothesis, the physical distance of each epileptogenic region identified by the VEP to all the EZs comprising the clinical hypothesis was small (mean: 5.67 mm; see Fig. 6**c**).

**Fig. 6.**
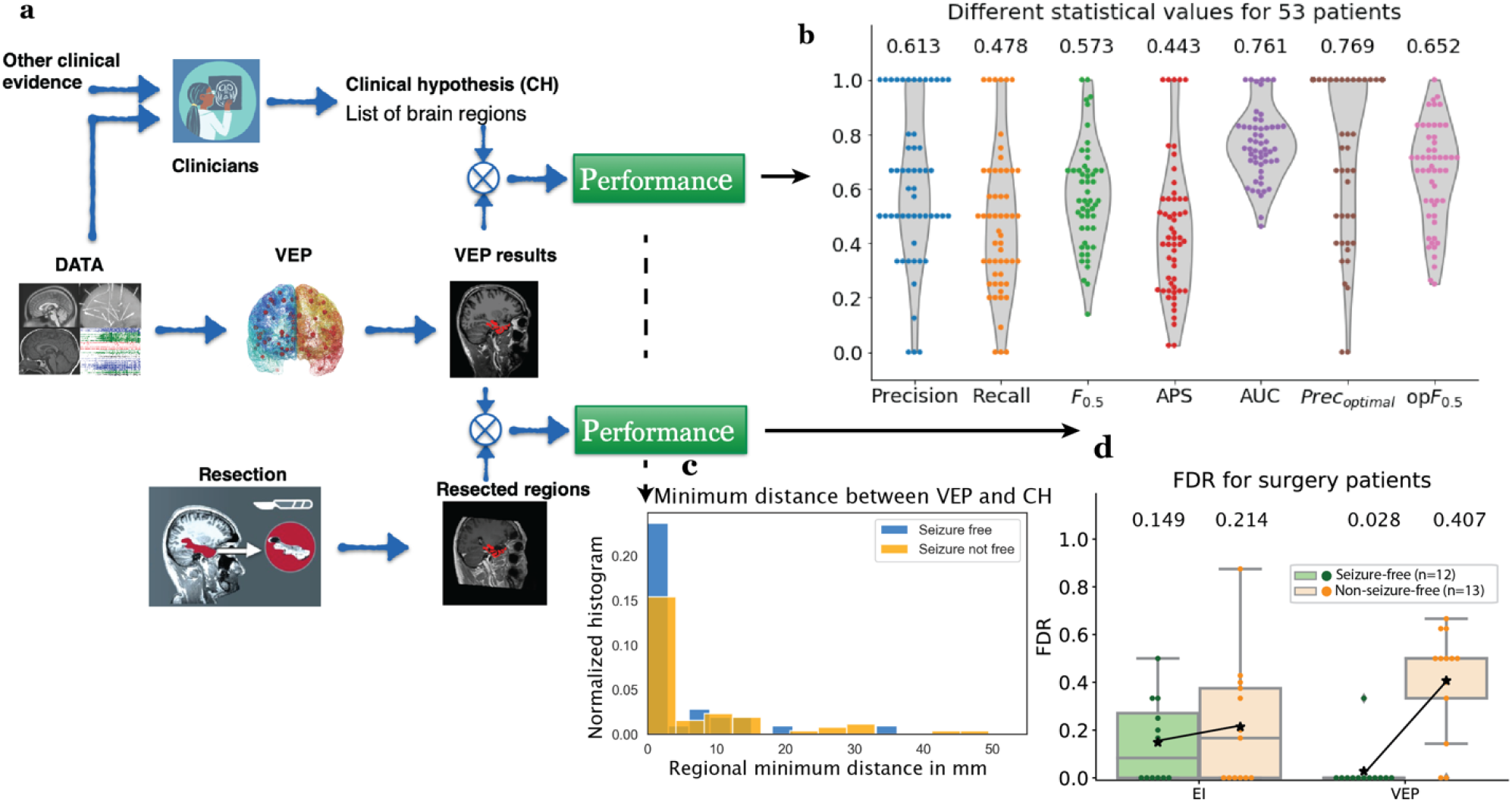
Performance of VEP. **a**. The evaluation framework includes two blocks. Top, the VEP results were evaluated against the clinical hypothesis based on a presurgical assessment. Bottom, the VEP results were evaluated against resected brain regions in the post-operative MRI. **b**. The distribution of the seven statistical evaluation results for 53 patients: precision, recall, F_0.5_, APS, AUC, Prec_optimal_, and opF_0.5_. For each measure, a violin plot demonstrates the distribution of 53 patients with each dot representing a patient. Above each violin plot is the mean value of a statistical measure across the 53 patients. **c**. The histogram of the minimal distance between each identified VEP region and the clinical hypothesis in the two groups in terms of surgical outcome, that is, seizure-free vs. not-free-seizure. **d**. False discovery rate (FDR) of the Epileptogenicity Index (EI) and VEP against post-operative MRI for 12 seizure-free patients and 13 non-seizure-free patients. Above each box plot is the mean value of the FDR.

When the VEP predictions were compared to the resected brain regions, we used the false discovery rate (FDR) as a performance metric in which false positives become more important. With seizure-free patients, a false positive estimate has a high possibility of being a true false EZ, whereas in not-seizure-free patients, the false positive estimate has a high possibility of indicating that the model is missing regions that would cause the seizures to remain. The VEP showed a very small false discovery rate (FDR) (mean: 0.028) for seizure-free patients (see **Fig. 6e**) compared with the EI (where we simply projected the EI results automatically to the VEP regions, mean: 0.149). For the not-seizure-free group, the VEP results showed a large increase in the FDR (mean: 0.407) compared with the seizure-free patients, suggesting that it may be possible to better exploit the predictive power of the VEP. Please note that the large resection regions in the current surgery strategy in some patients also contributed to the low FDR value to a certain degree.

## Discussion

VEP is a multimodal probabilistic framework for a personalized end-to-end analysis of brain imaging data from drug resistant epilepsy patients. The VEP workflow provides all necessary and optimized modules using the NMMs to estimate the EZN and provide a virtual surgery strategy in drug resistant epilepsy. All the modules are state-of-art and well-tested. The VEP workflow provides a successful use of brain models, compared with traditional spectral analysis of SEEG signals, such as the EI^3^, cEI^4^, epileptogenicity map^5^, and EZ fingerprint^7^. Compared with other model-based methods, such as neural fragility^33^ and dynamic network biomarker^34^ based on stability concepts in complex systems theory^16^, the VEP provides the first fully nonlinear system analysis of whole brain NMM and works on the whole brain source spaces instead of the sensor recording spaces alone. In this paper, we have provided a clear definition of the EZN from a modeling perspective and the corresponding calculations. Both the full sampling of the HMC in the Bayesian inference and the neural field high resolution simulation are state of the art. The current VEP has been extensively tested using a cohort of 53 retrospective patients and is now being evaluated prospectively in a large French clinical trial (EPINOV NCT03643016) recruiting 350 patients from 11 epilepsy centers. The main objective of EPINOV is to evaluate the capacity of the Virtual Epileptic Patient (VEP) algorithms to improve clinical outcomes in patients suffering from focal drug-resistant epilepsy undergoing SEEG surgery.

Other studies have shown that the EZN is highly correlated with the brain structural lesions observed in MRI^35^. In addition, certain neurotransmitters and glucose showed hypometabolism in the EZN, as shown by PET studies^36^. These additional pieces of prior knowledge could be integrated into the VEP workflow by defining the prior distribution. Dynamic causal modelling (DCM) is a general framework that allows for the analysis and variational Bayesian inversion of NMMs^37^. Using DCM, two related studies investigated the fluctuation of synaptic parameters^38^ or synaptic coupling weights^39^ in order to understand the generation of seizure activities. Compared to VEP, the studies utilizing DCM were based on a small number of cortical regions, and the non-linear NMM was approximated by its linearization. Recent studies have demonstrated the advantages of brain modeling in aiding epilepsy surgery by using the personalized information and by capably comparing different strategies in silico^18,20,21,40–42^. The virtual surgery module in the VEP workflow provides an all-in-one solution, which uses the generative function of large-scale brain modeling in TVB, a personalized connectome, and a patient-specific estimate of the EZN and its parameters from the VEP diagnostic modules.

Neural mass modeling approaches reduce thousands of vertices of source activity into one node mapped to a VEP region. Any grouping cancels out the directionality of the current dipole of the folded cortical sheet, which may lead to wrong mapping from sources to sensors and thus possibly introduce errors into the estimation of the EZN. The solution is to use model inversion on the high resolution of NFM. The huge dimensions are a big challenge for any inference method and needs to be overcome in the future. Meanwhile, we are systematically assessing the ability of the current VEP pipeline of the NMM with the synthetic data of NFM to try to answer two fundamental questions: whether the neural mass version of VEP pipeline is good enough to identify the ground truth, which was used in the generated data from the NFM, and under which conditions the VEP in the NMM version would succeed or fail.

Introducing regional variance may provide another fundamental way to improve the VEP pipeline. Currently, identical parametrization, with the exception of the excitability parameter related to seizure generation, is assumed across all sites of the brain. The connectivity network is another element that can influence the regional variance because of different connection strengths. However, the rich repertoire of anatomical data shows differences in cell type, cell architecture, receptor density, and functional specialization between brain areas^43,44^. Some subject-specific 23Na imaging data indicated different levels of excitability of different brain areas, which could be used to construct more subject-specific models ^45^. This region variance has been also demonstrated using the power spectra and peak frequencies of functional data, such as SEEG^22,46^. The big challenge is to find a meaningful mapping from the empirical data to the parameters in the neural model. In addition, because most anatomical and functional data about regional variance are available only on the group level, the balance between group and personalized information remains an important issue. All these further adventures in high resolution and regional variance are based on the solid VEP workflow we introduced in this paper.

## Data Availability

An example dataset used in the paper can be accessed at: https://doi.org/10.5281/zenodo.5816706. This data includes a patient's preprocessed anatomical and high-resolution functional simulation data. The patient raw datasets cannot be made publicly available due to the data protection concerns regarding potentially identifying and sensitive patient information. Interested researchers may access the data sets by contacting the corresponding authors.

## Methods

### Patient data

We obtained the dataset from 53 patients with drug resistant focal epilepsy who underwent a standard presurgical protocol at La Timone hospital in Marseille. Informed written consent was obtained for all patients in compliance with the ethical requirements of the Declaration of Helsinki and the study protocol was approved by the local Ethics Committee (Comité de Protection des Personnes sud Méditerranée 1). The non-invasive evaluation included assessing the patients’ clinical records, neurological examinations, neuropsychological testing, and EEG recordings. The subjects’ clinical data are given in Supplementary Table 1. Fifty-three patients (25 males, 28 females) were included. Mean age at epilepsy onset was 16.2 years (range 0.1-55), mean duration of epilepsy was 17.2 years (range 3-45), mean age at evaluation: 33.4 years (range 15 – 60). The evaluation included non-invasive T1-weighted imaging (MPRAGE sequence, repetition time = 1.9 or 2.3 s, echo time = 2.19 or 2.98 ms, voxel size 1.0 × 1.0 × 1.0 mm^3^) and diffusion MRI images (DTI-MR sequence, either with an angular gradient set of 64 directions, repetition time = 10.7 s, echo time = 95 ms, voxel size 1.95 × 1.95 × 2.0 mm^3^, b-weighting of 1000 s × mm^2^, or with an angular gradient set of 200 directions, repetition time = 3 s, echo time = 88 ms, voxel size 2.0 × 2.0 × 2.0 mm, b-weighting of 1800 s/mm2. The images were acquired on a Siemens Magnetom Verio 3T MR-scanner. All patients had invasive recordings stereo-EEG (SEEG) obtained by implanting multiple depth electrodes, each containing 10–18 contacts 2 mm long and separated by 1.5- or 5-mm gaps. The SEEG signals were acquired on a 128 channel Deltamed/Natus system with at least a 512 Hz sampling rate and recorded on a hard disk (16 bits/sample) using no digital filter. Two hardware filters were present in the acquisition procedure: a high-pass filter (cut-off frequency equal to 0.16 Hz at -3 dB), and an anti-aliasing low-pass filter (cut-off at one third of the respective sampling frequency). After the electrode implantation, a cranial CT scan was performed to obtain the location of the implanted electrodes.

### Data processing

To construct the individual brain network models, we first preprocessed the T1-MRI and DW-MRI data. Volumetric segmentation and cortical surface reconstruction were from the patient-specific T1-MRI data using the recon-all pipeline of the FreeSurfer software package (*http://surfer.nmr.mgh.harvard.edu*). The cortical surface was parcellated according to the VEP atlas^22^, and the code is available at *https://github.com/HuifangWang/VEP_atlas_shared.git*. We used the MRtrix software package to process the DW-MRI ^47^, using the iterative algorithm described in^48^ to estimate the response functions and subsequently used constrained spherical deconvolution^49^ to derive the fiber orientation distribution functions. The iFOD2 algorithm^50^ was used to sample 15 million tracts. The structural connectome was constructed by assigning and counting the streamlines to and from each VEP brain region. The diagonal entries of the connectome matrix were set to 0 to exclude self-connections within areas and the matrix was normalized so that the maximum value was equal to one. We obtained the location of the SEEG contacts from post-implantation CT scans using GARDEL (Graphical user interface for Automatic Registration and Depth Electrodes Localization), as part of the EpiTools software package^51^. Then we coregistered the contact positions from the CT scan space to the T1-MRI scan space of each patient.

### VEP model construction

We used virtual whole brain neural models^8^ to predict individualized EZs. These models represent the brain areas as nodes, which are connected through edges formed by white matter fibers. The activity of each node was modeled with a set of dynamical equations. The connection strength between nodes was inferred from the structural connectome derived from DW-MRI data. To model seizure-like activity we used the phenomenological 6D Epileptor model^23^, which consists out of 3 neural populations and acts on fast, intermediate, and slow time scales.

The Epileptor model consists of 6 coupled differential equations.

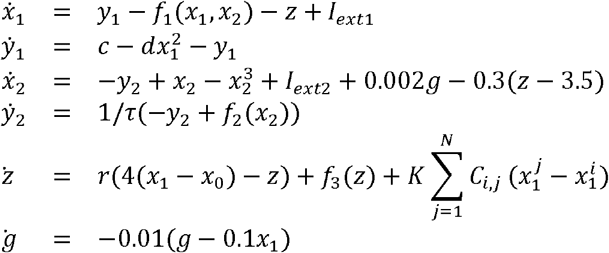

where

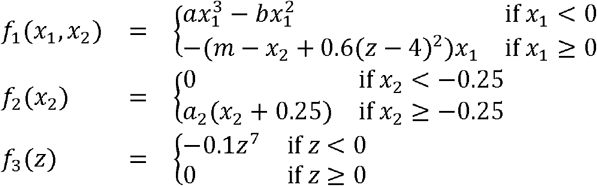

The state variables *x*_1_ and *y*_1_ describe the activity of neural populations on a fast time scale and can model fast discharges. The state variables *x*_2_ and *y*_2_ operate on an intermediate time scale and describe spike wave events. The oscillation of the slow permittivity variable *z* drives the system autonomously between ictal and inter-ictal states. The parameter *x*_0_ indicates the degree of excitability and directly controls the dynamics of the neural population to produce the seizure or not. The state variable *g* acts as a low-pass filter of the coupling from *x*_1_ to *x*_2_. The default parameters are *I*_*ext*1_*=* 3.1, *I*_*ext*2_*=* 0.45,*c* = 1, *d* = 5,*τ* = 10, *r* = 1,*K* = − 5,*a*= 1,*m* = 1, *a*_2_ = 6. The coupling between nodes of the network is defined by *C*_*i,j*_, which comes from the structural connectivity. *K* scales the connectivity, which can be varied between simulations to investigate different scenarios.

Taking advantage of time scale separation and using averaging methods, the 6D Epileptor is reduced to a 2D system^52^:

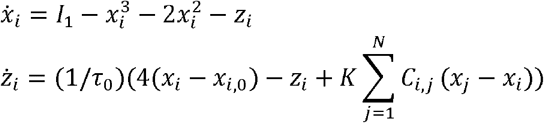

where *τ*_0_ scales the length of the seizure. The external input is defined as *I*_1_= 3.1. We used the 2D Epileptor for model inversion and estimated the parameters of the model from empirical data.

### Neural mass modeling (NMM) and neural field modeling (NFM)

Neural masses represent the macroscopic behavior of populations of neurons. They are defined by a set of differential equations that govern their dynamics. In TVB, neural masses represent the activity of a single brain area. Neural masses are linked through the structural connectome to form a full brain network model. The global equation for such a model can be given by

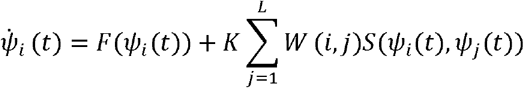

where *ψ*_*i*_ (*t*) is a state vector of neural activity at brain region *i* and time *t*. 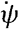 is the temporal derivative of the state vector. *F* is a function of the state and captures the local neural activity. In our case, *F* reflects the Epileptor model, described above. *W* is a matrix of heterogeneous connection strengths between node *i* and *j. S* is a coupling function of the local state *ψ*_*i*_ and the distant delayed state *ψ*_*j*_. That a node receives input through the network is given by the sum across the number of nodes *L* and scaled by a constant *K*. In this paper, this set of differential equations is solved using an Euler integration scheme with a step size of 0.5 ms in the virtual surgery simulations.

Neural fields extend neural masses along the spatial dimension. Instead of accumulating neural activity in a single point, the activity is represented continuously across the cortical surface. The global equations for such a model can be written as

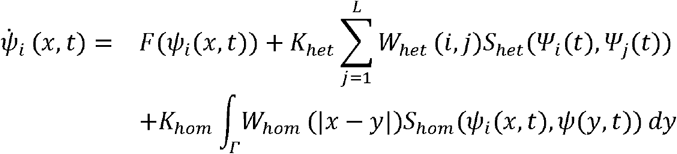

The spatial dimension of the field is given by 2 closed surfaces representing the pial cortex, each for one hemisphere. The surface is discretized by ∼260,000 vertices. The neural state of a vertex *i* at position *x* and time *t* is given by *ψ*_*i*_ (*x,t*). The subscript *i* indicates the brain region to which the vertex is assigned. There are 2 types of coupling in this model. The heterogeneous coupling through *W*_*het*_ and *S*_*het*_ is similar to the previous NMM approach, but the input to the coupling function *S*_*het*_ now is *ψ*_*i*_ and *ψ*_*j*_, the average neural state of all vertices belonging to regions *i* and *j*, respectively. The homogeneous coupling is given by the integral across the spatial domain *Γ. W*_*hom*_ is a translation-invariant coupling strength function based on the geodesic distance between vertices located at position *x* and *y* along the surface. In our implementation, this function is described by 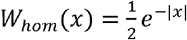. *S*_*hom*_ is coupling function of the neural state *ψ*_*i*_ at local position *x* and time *t* and the neural state *ψ* at position y and time *t*, regardless of the region assignment of the vertices. *K*_*het*_ and *K*_*hom*_ are the scaling parameters for heterogeneous and homogeneous couplings, respectively. Including both couplings with the increase in spatial resolution of the model from 162 brain regions in NMM to ∼260000 vertices in MFM causes greatly increased computational demand.

### Forward solution: NMM and NFM

Mapping the neural activity from the sources (VEP brain regions in NMM and vertices in NFM) to the sensors (SEEG contacts) is done by solving the forward problem and estimating a source-to-sensor matrix (Gain Matrix). As sources for our model, we used the vertices of the pial surface and volume bounding surfaces for the cortical and subcortical regions respectively. Surfaces are represented as triangular meshes. For the NMM approach we estimate that the matrix *g*_*j,k*_ from source brain region *j* to sensor *k* is equal to the sum of the inverse of the squared Euclidean distance *d*_*i,k*_ from vertex *i* to sensor *k*, weighted by the area *a*_*i*_ of the vertex on the surface.

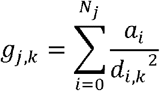

Here vertex *i* belongs to region *j* which has *N*_*j*_ vertices in total. The area *a*_*i*_ of vertex *i* is obtained by summing up one-third of the area of all the neighboring triangles. Vertices belonging to the same brain region are summed to obtain the gain for a single region of our brain network model. The resulting gain matrix has dimensions *M* × *N*, with *M* being the number of regions and *N* the number of sensors. Matrix multiplication of the simulated source activity with the gain matrix yields the simulated SEEG signals.

This distance-based approach and the summation of all vertices within each region neglects the orientation of the underlying current dipoles. Pyramidal neurons, which are oriented normal to the cortical surface, are assumed to be the physiological source of any electrical signal recorded with SEEG, EEG, or MEG^32^. This could not be taken into account when modeling a brain region by a NMM. However, approaches that use neural fields have this ability. To solve the forward problem we follow the analytical solution proposed in [24] for electrical fields in an unbounded homogeneous medium. We neglect possible boundary effects because SEEG records within the brain where electrical conductivity is almost constant. Therefore, we can estimate the gain matrix elements *g*_*i,k*_ for the NFM approach by

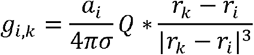

where *r*_*k*_ and *r*_*i*_ are the position vectors of sensor *k* and source vertex *i*, respectively. |*v*| represents the L2 norm of a vector *v. Q* is the dipole orientation vector and *σ* the electrical conductivity. Since we assume constant conductivity across the brain, it becomes merely a scaling factor, which we set to *σ =* 1.

### Estimation of a prior

The model inversion module needs priors, which can be either uninformative or informative. For uninformative priors, we gave all the brain regions with the same distributions, such as normal distributions, for fitting parameters. The mean of the normal disturb for the excitability parameter *x*_0_ = -3.0, which means that the prior assumes that all brain regions are highly possible in the healthy zones. For the informative prior, we would like to give a bias to some regions with a higher excitability parameter *x*_0_, i.e., prior knowledge indicates that these selected regions have higher possibility of being in the EZN. The selection of these regions may come from expert knowledge, such as MRI lesion information, or from a clinical hypothesis. Here we designed an algorithm to estimate the prior using the signal processing of the recorded raw seizure SEEG data. We compute the spectrogram of the time series using Fourier transforms in consecutive windows, with a window length of 1 second if the seizure duration is longer than 200 seconds, otherwise, an overlap of 1 s windows is used, such that in total 200 windows are calculated. Values of the spectrogram smaller than 10^−15^ are set to 10^−15^ and all values are log transformed. A sensor-prior vector, which identifies the seizure onset in each SEEG channel, is calculated based on the channels’ spectral content across 52 different frequency bands. The metric is computed across two nested loops. In the outer loop we increase the lower-bound frequency from 10 Hz to 90 Hz in steps of 10 Hz. In the inner loop we increase the upper-bound frequency from the current lower-bound + 10 Hz to 120 Hz in steps of 10 Hz. The frequency band used in each run of the inner loop is defined by the range from the lower-to the upper-bound frequency. Next, we average the spectrogram across the defined frequency-band, resulting in one average time series per SEEG channel. This time series is thresholded by its 90% percentile. Non-0 values are set to 0 if fewer than 5 consecutive time points are above the threshold.

After thresholding, we find the first time point above the threshold in each channel. To assign high values to channels with early onset, we use the reciprocal of this time point. Reciprocal values are normalized so that the maximum value equals one. High values of this metric indicate an early increase in the specific frequency band that can be expected to indicate the seizure onset in a specific SEEG channel. All the above computations are performed on the sensor level. Thus, we use a sensor-to-source-matrix based on the distance from the SEEG contacts to the vertices of the brain regions to assign values to each brain region. The entries of this matrix are 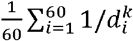, where 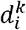 is the Euclidean distance from sensor *k* to source vertex *i*. We consider the 60 closest vertices of each region to the sensor, to avoid bias coming from variability in the size and shape of regions, i.e., when considering all vertices of a region in this computation, smaller regions could dominate over bigger regions because the mean across all vertices of the bigger region could be smaller than that of the smaller region, even in situations in which the bigger region is the closer one to the sensor. On the other hand, if we only considered the few closest vertices of each region, the method would be too sensitive, potentially mislabeling several vertices in the automatic parcellation.

We proposed two methods for projecting the values to the source level. For method VEP-M (M is for maximum) we find the brain region in the sensor-to-source-matrix that projects the strongest to a particular sensor and assign the sensor-prior value to that region. If a region projects strongest to multiple sensors, the highest value across the sensors is assigned to the region. For the VEP-W method (W is for weighted), we multiply the sensor-prior vector by the sensor-to-source-matrix to project to the brain regions. This way one onset value per region is calculated for each frequency band. To get one final prior epileptogenicity parameter per region, we average the onset values across all the frequency bands and divide by the maximum across the regions. Those brain regions with a resulting value above a threshold of 0.5 are assigned a mean of the normal distribution for the epileptogenicity parameter *x*_0_ of -1.5; all other regions are assigned a value of -3.0 as the mean. The threshold can be adjusted manually in some special cases, such as if different patterns need to be tested. Channels with high amplitude physiological activity might impact the prior estimation; thus, we suggest manually removing them.

### The optimization MAP pipeline

We apply a Bayesian modeling approach to infer the epileptogenicity parameters and source time series for each seizure. According to Bayes’ theorem, the posterior probability distribution *p*(*θ*|*y*) of a parameter *θ* given the data *y* is equal to the product of the likelihood *L*(*y*|*θ*) of the data given the parameter and the prior probability distribution *p*(*θ*) of the parameter divided by the marginal likelihood *p*(*y*) of the data.

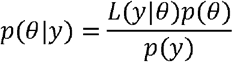

The marginal likelihood *p*(*y*) = ∫*L* (*y*| *θ*) *p*(*θ*)*dy* is intractable to compute in such complicated multivariate models as VEP. Since the marginal likelihood is a normalizing constant to scale the integral across the posterior distribution to 1, one can state the Bayes theorem as

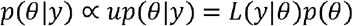

Where the posterior probability is proportional to the unnormalized posterior probability *up* (*θ*|*y*). Normalized and unnormalized posterior distributions have the same shape, maxima, and minima; they are just scaled by constant versions of each other. Markov chain Monte Carlo methods generate samples of the unnormalized posterior distribution. Given enough samples the posterior distribution can be approximated and inference can be drawn about the parameters of interest. In the optimization pipeline, we obtain the maximum-a-posteriori estimate *θ*_*MAP*_ using the L-BFGS quasi-Newton method.

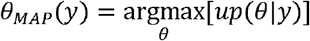

To find *θ*_*MAP*,_ the optimization algorithm performs an iterative process, starting with an initial guess of the parameters and then moving through parameter space following the direction of the gradient of the probability distribution. The algorithm terminates after either a maximum of 20,000 steps or convergence has been reached. Convergence is identified when changes in parameters, gradients, or probability density between steps are below a certain threshold. In the case of the VEP model, the posterior probability is given by the product of the prior probabilities of each parameter and the likelihood of the data. Stan transforms probabilities into log probabilities, such that products of probabilities become sums. The log of the posterior probability is then the sum of the log likelihood and the log prior probabilities of all the parameters. We specified the prior probabilities for the epileptogenicity parameter *x*_*i*,0_ for brain region *i*, the time scale of the slow variable *τ*_0_, one scaling *s* and one additive *a* constant of the simulated SEEG, the global coupling scaling factor *K*, and the initial conditions for state variables *x*_*i*_(*t*_0_) and *z*_*i*_ (*t*_0_) in region *i*, as well as the distribution width of the extracted data features *ϵ*_*v*_.

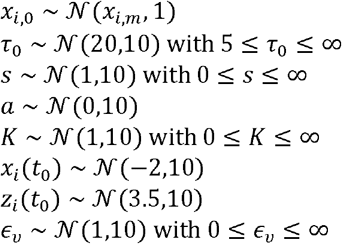

where 𝒩(*μ, σ*) is a normal distribution with mean *μ* and dispersion *σ* and *x*_*i,m*_ ∈ {−3, −1.5} is the epileptogenicity prior for each brain region. Some prior probabilities are truncated by setting a possible minimum value. The likelihood function is given by

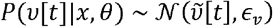

where *v* and 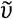 are the empirical and estimated SEEG data features. In our case, we considered the data feature to be both the seizure envelopes and the simulated SEEG channel power. Two algorithmic diagnostic metrices are goodness of convergence and goodness of fit. Goodness of convergence represents the number of runs that terminated properly, i.e., the varying of the likelihood converges to the given threshold. Goodness of fit is defined by 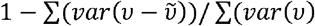, where the sum is across all the SEEG recorded channels.

### The sampling HMC pipeline

In the sampling pipeline, we applied the No-U-Turn Sampler (NUTS), a self-turning variant of the HMC algorithm to sample the posterior density *p*(*θ*|*y*) of the model parameters. The performance of the HMC is highly sensitive to step size and number of steps in the leapfrog integrator for updating the position and momentum variables in a Hamiltonian dynamic simulation^53^. We use NUTS, which is implemented in Stan and which extends HMC with adaptive tuning of both the step size and the number of steps in a leapfrog integration to sample efficiently from the posterior distributions^53^,^27^. To overcome the inefficiency in the exploration of the posterior distribution of the model parameters, we used a reparameterization of the model parameters based on the map function from the model configuration space to the observed measurements. Our reparameterization-based approach significantly reduces the computation time by providing higher effective sample sizes and removing divergences by exploring the posterior distributions of the linear combinations of regional parameters that represent the eigenvectors obtained from the singular value decomposition of the gain matrix. We denote the matrix of the eigenvectors of *G*^*T*^*G* as *V* and the new parameters as 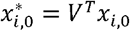 and 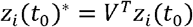.

We assess model identifiability based on analysis of posterior samples, which demonstrates that the sampler explores all the modes in the parameter space efficiently. The analysis includes trace-plots (evolution of parameter estimates from draws over the iterations), pair plots (to identify collinearity between variables), and autocorrelation plots (to measure the degree of correlation between draws of samples). Sampling convergence of the algorithms is assessed by estimating the potential scale reduction factor and calculating the effective sample size based on the samples of the posterior model probabilities, providing estimates of the efficient run times of the algorithm.

### Calculation of the epileptogenic values

After the optimization and sampling pipeline, we obtained the estimated source time series, and based on them we calculated brain region-specific epileptogenicity values (EVs). The source time series (variable “x” of the 2D Epileptor) of region *i* is checked for values above a threshold. This threshold is set to 0 for empirical data. The first occurrence of such a value is considered to be the onset of the seizure *t*_*i*_ in that region. We set *t*_0_= *min*(*t*_*i*_),*i* = 1,…,162. Brain regions with no estimated seizure (i.e., no values above 0) are assigned an onset value *t*_*i*_ = 200. We calculate the *EV*_*i*_ of brain region *i* by 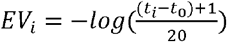. Then we normalize the *EV* vector to [0,1] for each run of either the optimization or the sampling pipeline. In this way, we obtain the distribution of EVs from the optimization pipeline while considering the sensitivity of the sensor spatial sampling. The sampling pipeline outputs the posterior distribution of the EVs.

### Sensor sensitivity and clinical reports

In the optimization pipeline, we performed bootstrapping to derive a measure of confidence for our estimated EVs, in terms of the robustness of the identified brain regions with respect to sensor location sensitivity. In each bootstrapping sample, a randomly chosen bipolar SEEG contact is removed from the full set and the remaining data is used to run the optimization algorithm in Stan. This means that the empirical data, which needs to be fitted in each run, contains one less SEEG envelope timeseries, one less SEEG estimated power, and one less row in the source-sensor mapping matrix. This procedure is repeated 100 times, selecting SEEG contacts randomly with replacement. All EVs are normalized by subtracting the minimum median EV and dividing by the difference between the maximum and minimum median EVs.

Those regions with a normalized median EV greater than a threshold of 0.5/3 are presented in the clinical report, beginning with the region with the highest normalized median EV. The clinical report contains 5 columns. From left to right, these columns contain the region name, the normalized median, the “confidence1” (i.e., percentage of normalized EVs above 0.5), “confidence2” (i.e., (1-std(EV)), as well as a violin plot in the right-most column, which depicts the distribution of the normalized EV values. The yellow dot in the middle of the distribution represents the normalized median EV.

### Integration algorithm for seizures and pipelines

Depending on the spatiotemporal dynamics patterns, the clinicians defined the different seizure types. These different seizure types may lead to different EZNs (2 out of 53 analyzed patients). We designed an algorithm to integrate the different runs on the different seizures belonging to the same seizure type for one patient. This algorithm can be used to integrate any combination of different runs, depending on the scientific questions being considered. In this paper, specifically for the evaluation module, we focused this integration procedure on two runs of the VEP prior algorithm, VEP_M and VEP_W, in the optimization pipeline. The reason is that these two pipelines can be automatically calculated without necessitating rich clinical knowledge, so it can be widely used.

For each seizure, the algorithm chooses VEP_M and VEP_W based on the variance of their bootstrapped EV values and the convergence of the optimization based on the likelihood values. Then we calculated the weights based on the variance of 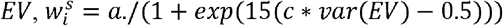, for brain region *i* and seizure *s*, with *a* = 1 and *c* = 3 as default values. Then we defined the *EV*^*m*^ for each region *i* by integrating the results from *S* multiple seizures from multiple pipelines, 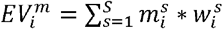, where *m*_*i*_ is the median values of EV on region *i* for seizure *s*.

### Performance measures

We used seven performance measures to compare the results against a given standard. Their calculation is based on four possible classification results: true positive *tp*, true negative *tn*, false positive *fp*, and false negative *fn*, where *p* and *n* are the number of positive and negative cases in the standard set and *t* and *f* respectively indicate whether the compared group agrees or disagrees with the standard groups. The detailed definition is shown in Supplementary Table 2. Precision reflects the positive predictive value, which expresses the probability of a given positive test result being truly positive. Recall (also called sensitivity) reflects the true positive rate, i.e., the proportion of correctly identified true positives to all positives. *F*_0.5_ is the harmonic mean of the precision and recall. In our cases, we put more emphasis on precision than on recall, because both the region lists from the clinician hypothesis and the post-operative MRI tended to provide more brain regions than the results of the VEP, which implies that false positive results are a more important measure than false negative ones. *APS* is another measure that combines precision and recall as the weighted mean of the precision achieved at each threshold *θ* with the increase in recall from the previous threshold used as the weight. The AUC is the area under the ROC curve, which is a plot of the true positive rate against the false positive rate (*fp*/*fp* + *tn*) at varying thresholds. Both *APS* and *AUC* consider all possible threshold by definition, which means that they can be considered to be threshold-free measures. We reported the precision, recall, and *F*_0.5_ for all the patients using fixed thresholds. Patient-specific situations are also important factors to consider, so we also demonstrated the measure *precision* and *F*_0.5_ using patient-specific thresholds.

**Supplementary table 1:** Patient information. Abbreviations: AVM: arteriovenous malformation; DNET: dysembryoplastic neuroepithelial tumor; FCD: focal cortical dyplasia; HS: hippocampal sclerosis; HH: hypothalamic hamartoma; L: left; NA: not applicable; PMG,:polymicrogyria; PNH: periventricular nodular heterotopia; R: right; Eng.: Engel score; NS: number of seizures analyzed for each patient.

| ID | Sex | Epilepsy type | MRI | Histopathology | Side | Eng | NS |
| --- | --- | --- | --- | --- | --- | --- | --- |
| 1 | F | Temporo-insular | Normal | HS | R | II | 4 |
| 2 | F | Temporo-occipital | L temporo-occipital PNH | NA | L | NA | 3 |
| 3 | M | Temporo-frontal | R temporo-occipital scar | FCD1a | R | I | 3 |
| 4 | F | Temporal | R temporal mesial ganglioglioma | Ganglioglioma | R | I | 4 |
| 5 | M | Postcentral - superior parietal | L postcentral-parietal gyration asymmetry | NA | L | NA | 3 |
| 6 | M | Fronto-temporal | Normal | NA | L | NA | 3 |
| 7 | M | Temporal | Normal | Slight gliosis | R/L | III | 2 |
| 8 | F | Temporal | L amygdala enlargement | Slight gliosis | L | III | 4 |
| 9 | F | Bifocal: parietal mesial & temporo-basal | Unknown R parietal lesion | Rosenthal fibers; slight gliosis | R | III | 5 |
| 10 | F | Temporal | L HS | HS | L | I | 6 |
| 11 | F | Frontal | L frontal scar (abcess) | Gliosis | L | IV | 3 |
| 12 | F | Bilateral temporo-frontal | Bilateral hippocampal & amygdala T2-hypersignal | NA | R/L | NA | 2 |
| 13 | M | Frontal | Normal | Slight gliosis | L | I | 3 |
| 14 | F | Premotor | Normal | FCD2b | L | I | 6 |
| 15 | M | Temporal | R temporal PMG & multiple PNH | NA | R | NA | 2 |
| 16 | M | Temporo-operculo-fronto-parietal | R temporo-parieto-insular & L temporo-parietal necrosis | NA | R/L | NA | 4 |
| 17 | M | Temporal | L temporo-polar hypothyrophy and hippocampal sclerosis | HS; gliosis | L | I | 2 |
| 18 | M | Parieto-temporal | L Parieto-occipital necrosis (perinatal anoxia) | NA | L | NA | 2 |
| 19 | M | Temporo-insular | Normal | NA | L/R | NA | 5 |
| 20 | F | Temporal | Normal | HS | R | I | 3 |
| 21 | F | Occipital | Normal | FCD1c | L | II | 4 |
| 22 | F | Parietal | L parietal FCD | FCD2b | L | I | 5 |
| 23 | M | Temporal | Normal | Gliosis | L | III | 2 |
| 24 | M | Temporal | Normal | NA | R | NA | 3 |
| 25 | M | Insular | Normal | NA | L | I | 3 |
| 26 | F | Occipital | PNH | NA | R | NA | 3 |
| 27 | M | Frontal | R prefrontal gliotic scar (AVM) | Gliosis | R/L | II | 3 |
| 28 | F | Temporo-frontal | Anterior temporal necrosis | Gliosis | R | III | 3 |
| 29 | F | Bilateral temporal | Bilateral posterior PNH | NA | R/L | NA | 9 |
| 30 | M | Temporo-frontal | R Frontal FCD | FCD 2 | R | I | 1 |
| 31 | F | Occipital | Normal | NA | R | NA | 4 |
| 32 | F | Bilateral temporal with HH | L HH | NA | R/L | NA | 5 |
| 33 | F | Temporo-parieto-opercular | Normal | HS | R | IV | 10 |
| 34 | F | Temporal | Multiple R temporo-parietal PNH & temporal PMG | NA | R | NA | 2 |
| 35 | M | Bilateral occipito-temporal | R occipital mesial FCD | NA | R/L | NA | 4 |
| 36 | F | Temporo-insular | R temporal anterior resection cavity | Gliosis | R | IV | 4 |
| 37 | M | Temporal mesial | R temporo-polar & amygdala FCD, L post-chiasmal pilocytic astrocytoma | FCD 2b | R | III | 4 |
| 38 | M | Bilateral temporal | Normal | NA | L/R | NA | 8 |
| 39 | M | Temporo-frontal | R fronto-temporal necrosis (gunshot injury) | Gliosis | R | I | 3 |
| 40 | F | Temporal mesial | Hippocampal sclerosis | HS | L | II | 2 |
| 41 | M | Bilateral, temporal predominant | R perisylvian necrosis (perinatal stroke) | NA | R/L | NA | 4 |
| 42 | F | Temporal mesial | Bilateral hippocampal sclerosis | NA | L | NA | 6 |
| 43 | F | Premotor | R precentral FCD | NA | R | NA | 3 |
| 44 | M | Multifocal: parieto-operculo-premotor; temporal mesial | L hippocampal & amygdala T2 hypersignal | NA | L | NA | 2 |
| 45 | F | Temporal mesial | Normal | NA | L | IV | 4 |
| 46 | M | Insulo-parieto-premotor | Normal | NA | R | NA | 1 |
| 47 | M | Bilateral frontal | Normal | NA | R/L | NA | 2 |
| 48 | F | Premotor | R parietal DNET | NA | R | NA | 1 |
| 49 | F | Motor-opercular | R fronto-opercular resection cavity | NA | R | NA | 1 |
| 50 | M | Motor-premotor | L insulo-opercular necrosis (stroke) | NA | L | I | 3 |
| 51 | F | Temporal mesial | Normal | NA | L | I | 2 |
| 52 | F | Prefronto-insular | Normal | NA | L | III | 4 |
| 53 | M | Temporal | Normal | NA | L | I | 3 |

**Supplementary table 2:** Mathematic definition of measures.

| Measures | Definition | Threshold |
| --- | --- | --- |
| precision |  | Fixed |
| recall |  | Fixed |
|  |  | Fixed |
|  |  | Threshold-free |
|  |  | Threshold-free |
|  | , where | personalized |
|  |  | personalized |

## Code availability

The code used to run VEP workflow will be available at *https://github.com/HuifangWang/VEP_INS_workflow*.

## Acknowledgements

This work is funded through the European Union’s Horizon 2020 Framework Programme for Research and Innovation under the Specific Grant Agreement No. 945539 (Human Brain Project SGA3) and the French National Research Agency (ANR) as part of the second “Investissements d’Avenir” program (ANR-17-RHUS-0004, EPINOV).

## Author contributions

V.J., F.B. and M.G. conceived the study and V.J. designed and supervised the study. H.E.W. and J.S. performed the data analysis. H.E.W., P.T., J.L, J.J., A.N.V., V.S., and S.M.V. wrote the algorithm and software with input of M.H., M.W and B.D. F.B. and J.S. gave the clinical input. H.E.W, P.T. and V.J. wrote the paper with input from all authors.

## Competing interests

The authors declare no competing interests.

**Supplementary Fig. 1.**
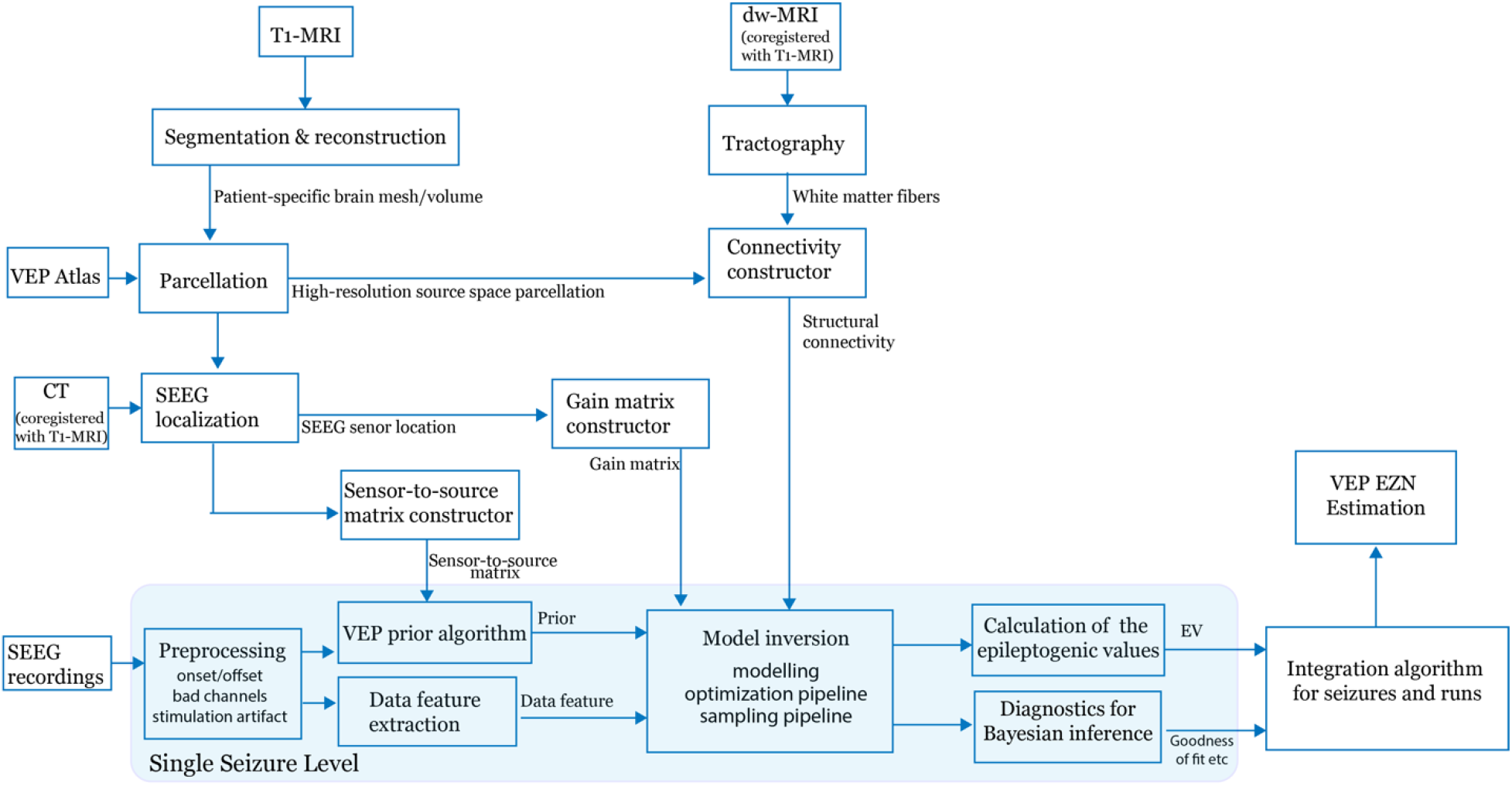
Flowchart of the VEP pipeline. First, T1-MRI defines the patient-specific high-resolution space and obtains the parcellation (brain regions on each vertex) according to the VEP atlas. Then we obtain the structural connectivity, gain matrix (source-to-sensor matrix), and sensor-to-source matrix in a patient-specific brain space by co-registering both dw-MRI and CT with T1-MRI. For each seizure, we run the VEP pipeline to obtain the EV distributions as well as the diagnostics, such as the algorithmic convergence and goodness of fit, for the MCMC and MAP algorithms. Then we can run the integration algorithm to combine the results for multiple seizures and runs and obtain the EZN estimate from the VEP pipeline.

**Supplementary Fig. 2.**
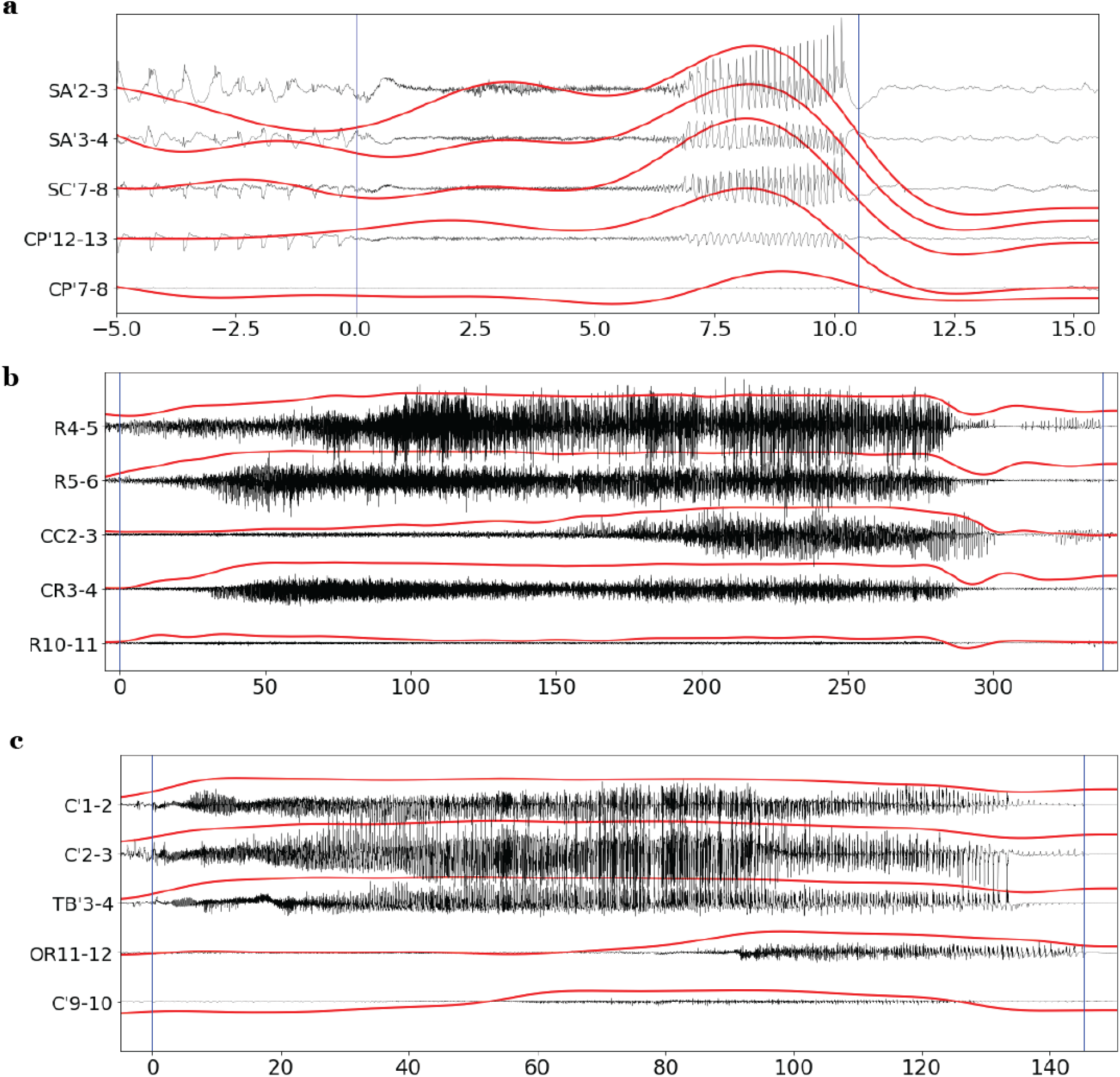
The data for three other seizure patterns from different patients. a, A short seizure with LVFA followed by periodic spikes. b, A long seizure with LVFA onset and then propagation to other regions with a similar amplitude. c, A long seizure with LVFA onset and then propagation to other regions with a lower amplitude.

**Supplementary Fig. 3.**
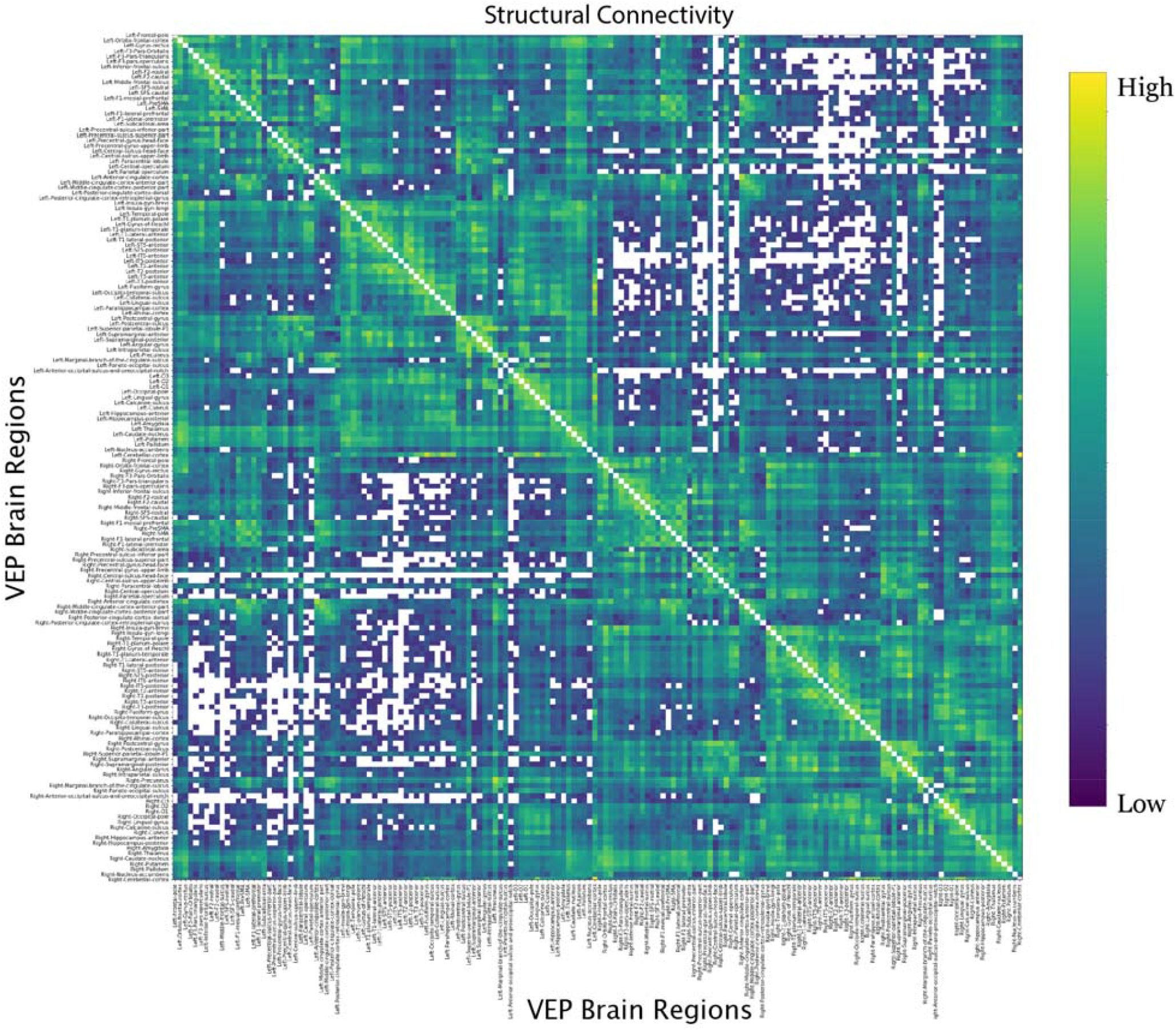
Structural connectivity between 162 VEP brain regions for the example patient.

**Supplementary Fig. 4.**
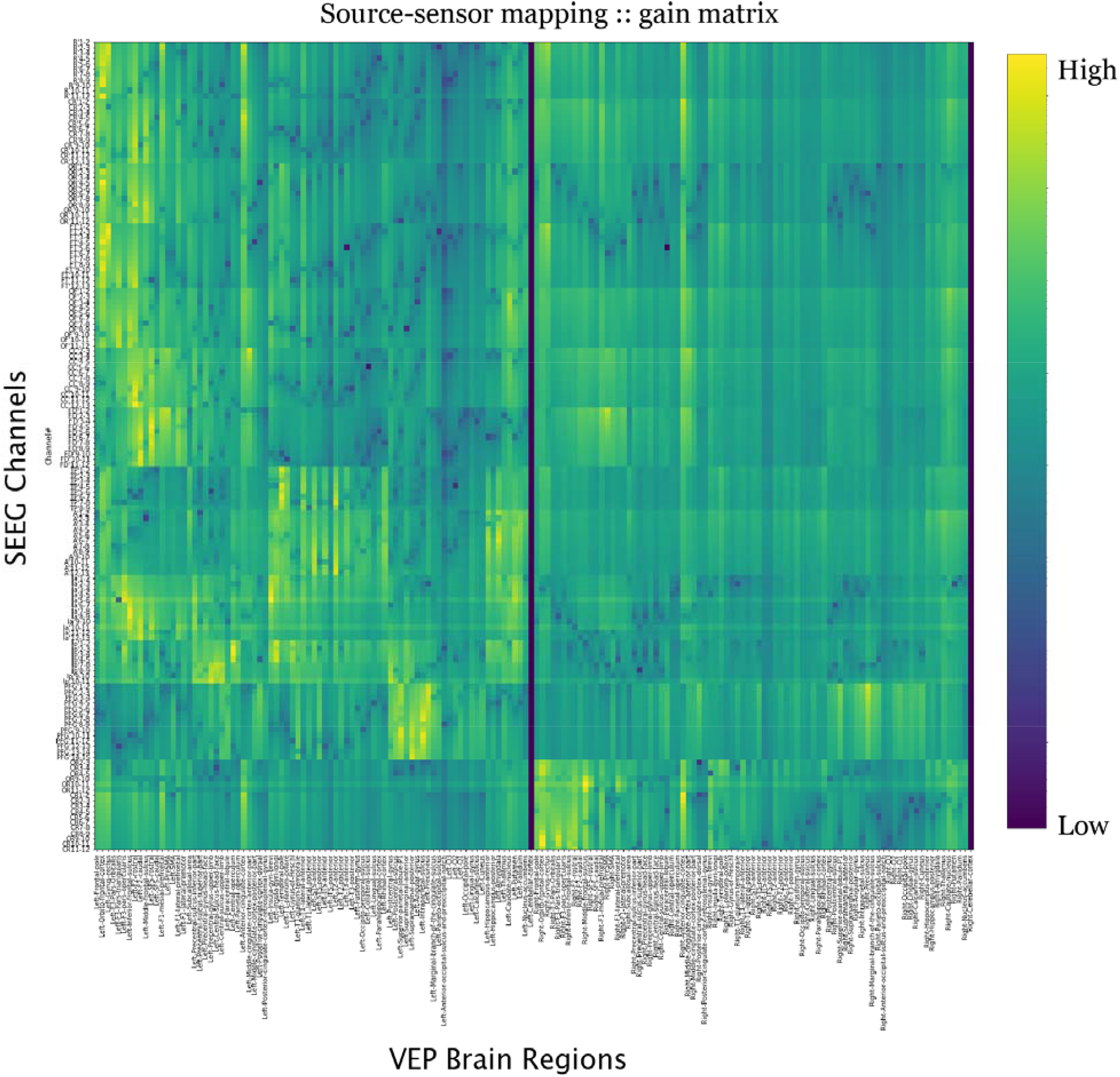
Source-to-sensor mapping (gain matrix) between recordings from SEEG bipolar electrodes and 162 VEP brain regions for the example patient.

**Supplementary Fig. 5.**
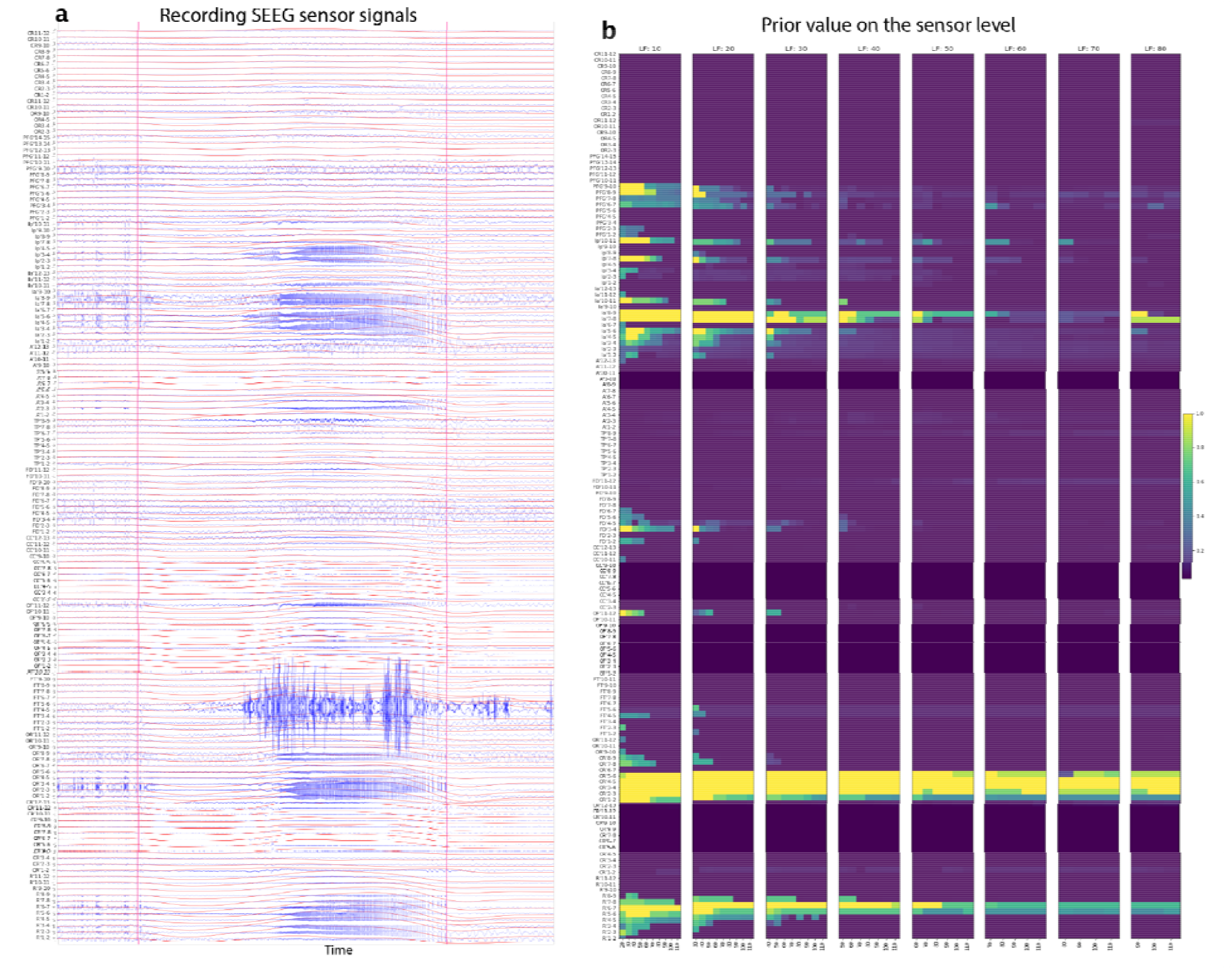
**a**, Raw time series of the SEEG sensor signals from the example patient (in blue) and the corresponding data feature (in red). **b**, The prior values of the sensors on 52 frequency bands; the low frequency boundary is from 10 Hz to 80 Hz with steps of 10 Hz, and the high frequency boundary is from 20 Hz to 110 Hz with steps of 10 Hz.

**Supplementary Fig. 6.**
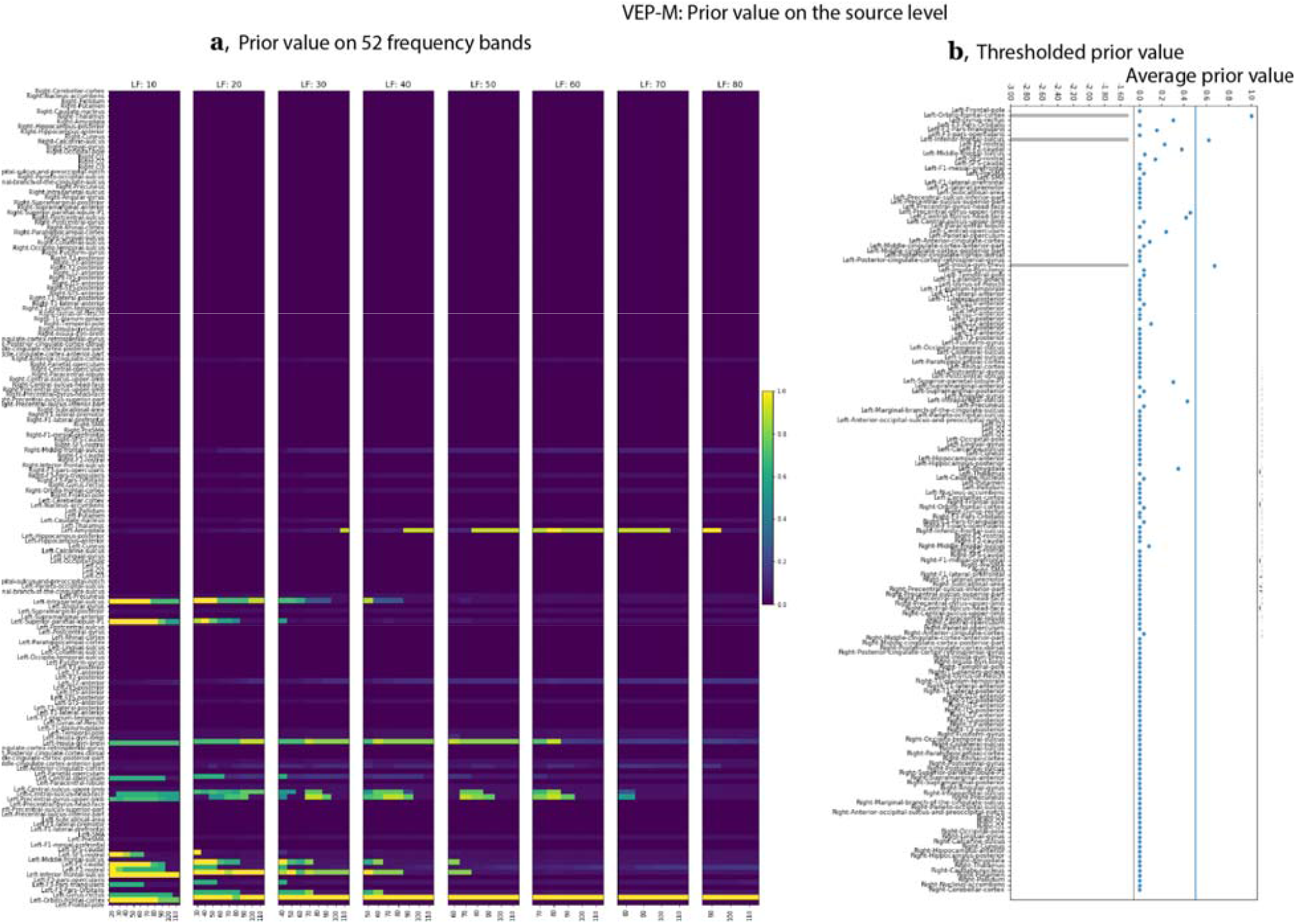
Prior value on the source level through the VEP-M algorithm. **a**. For each frequency band, the prior value VEP-M maps the prior value of the sensors that have the minimal distance to a given source. We considered 52 frequency bands: The low frequency boundary was from 10 Hz to 80 Hz with steps of 10 Hz, and the high frequency boundary was from 20Hz to 110 Hz with steps of 10 Hz. **b**, Right, the average across 52 frequency bands of prior values on brain regions. Left, the bar value of the prior values, which were used in the model inversion module for the VEP-M run. The threshold was 0.5.

**Supplementary Fig. 7.**
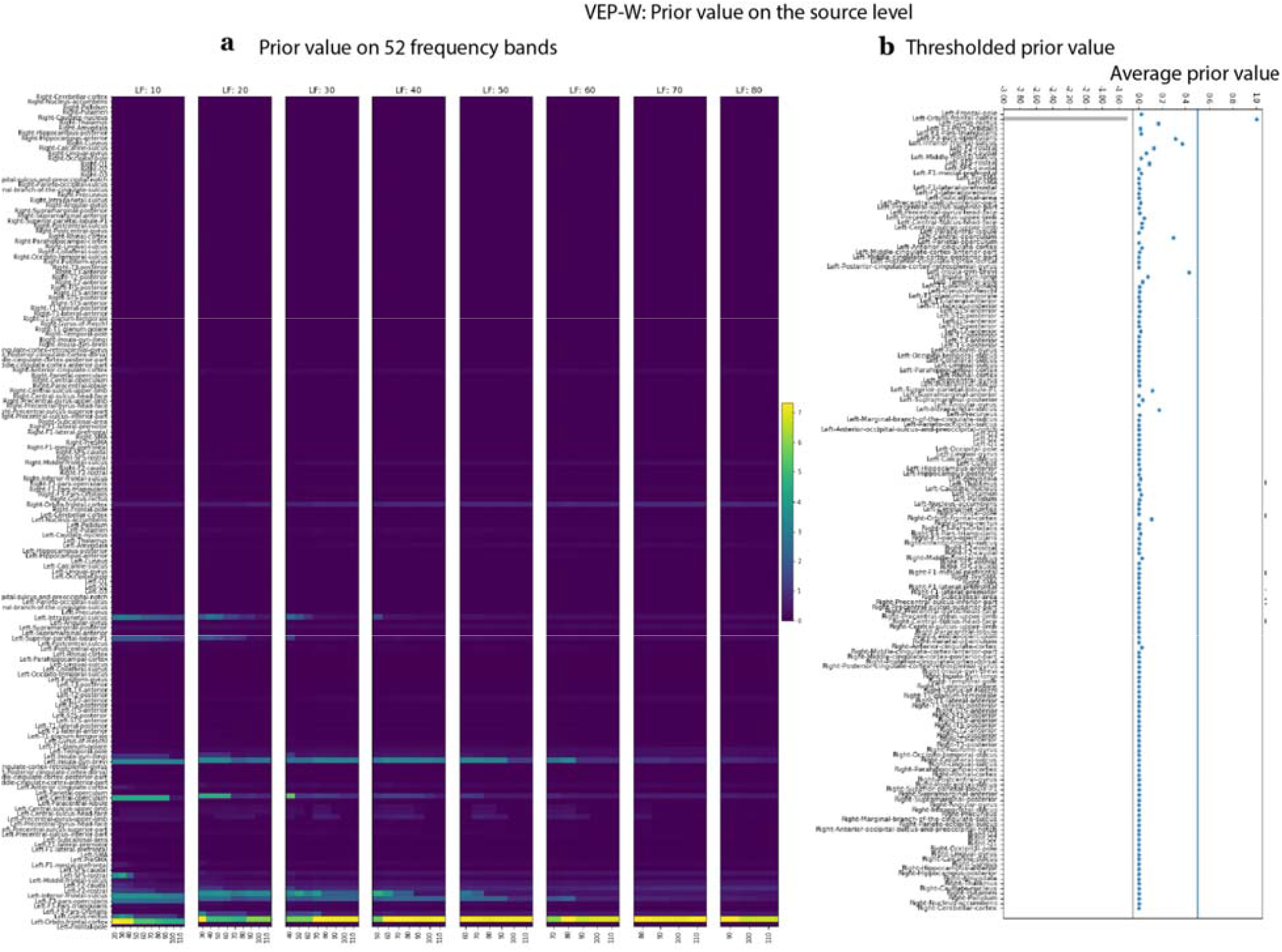
Prior value on the source level through the VEP-W algorithm. a. For each frequency band, the prior value VEP-W maps a weighted sum of the prior value of all sensors to each source. The weight is based on the distance between the source and the sensors. We considered 52 frequency bands: The low frequency boundary was from 10 Hz to 80 Hz with steps of 10 Hz, and the high frequency boundary was from 20 Hz to 110 Hz with steps of 10 Hz. b, Right, the average across 52 frequency bands of prior values on brain regions. Left, the bar value of the prior values that were used in the model inversion module for the VEP-W run. The threshold was 0.5.

**Supplementary Fig. 8.**
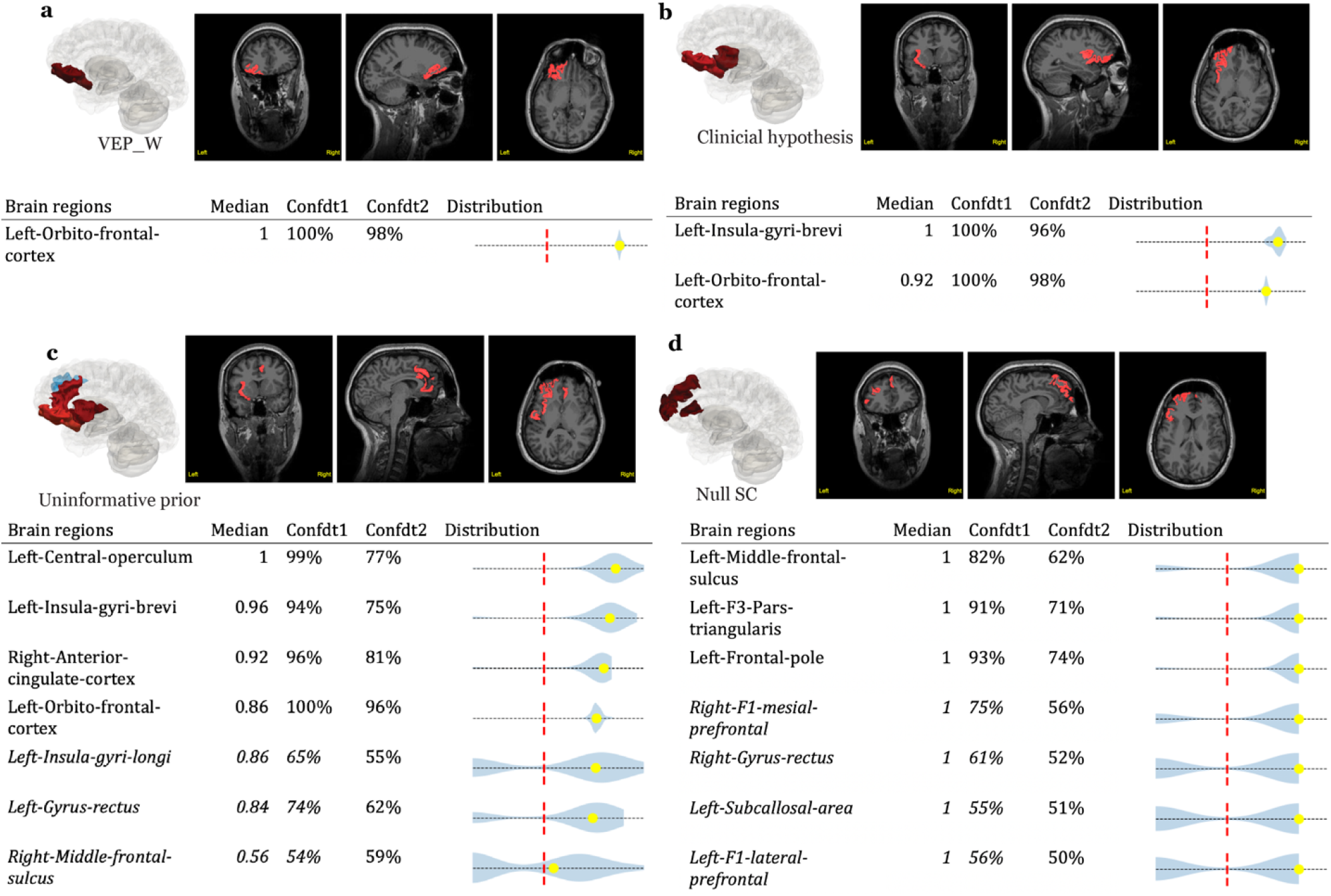
The results of 4 VEP runs in an optimization pipeline for the example patient. **a**, The results of a run using the VEP-W prior. **b**, The results of a run using the clinical hypothesis as the prior. **c**, The results of a run using an uninformative prior. **d**, The results of a run using an uninformative prior and no SC. Inside each subfigure: Top: the VEP results labeled in 3D mesh and MRI slides. Bottom, the clinical tables.

**Supplementary Fig. 9.**
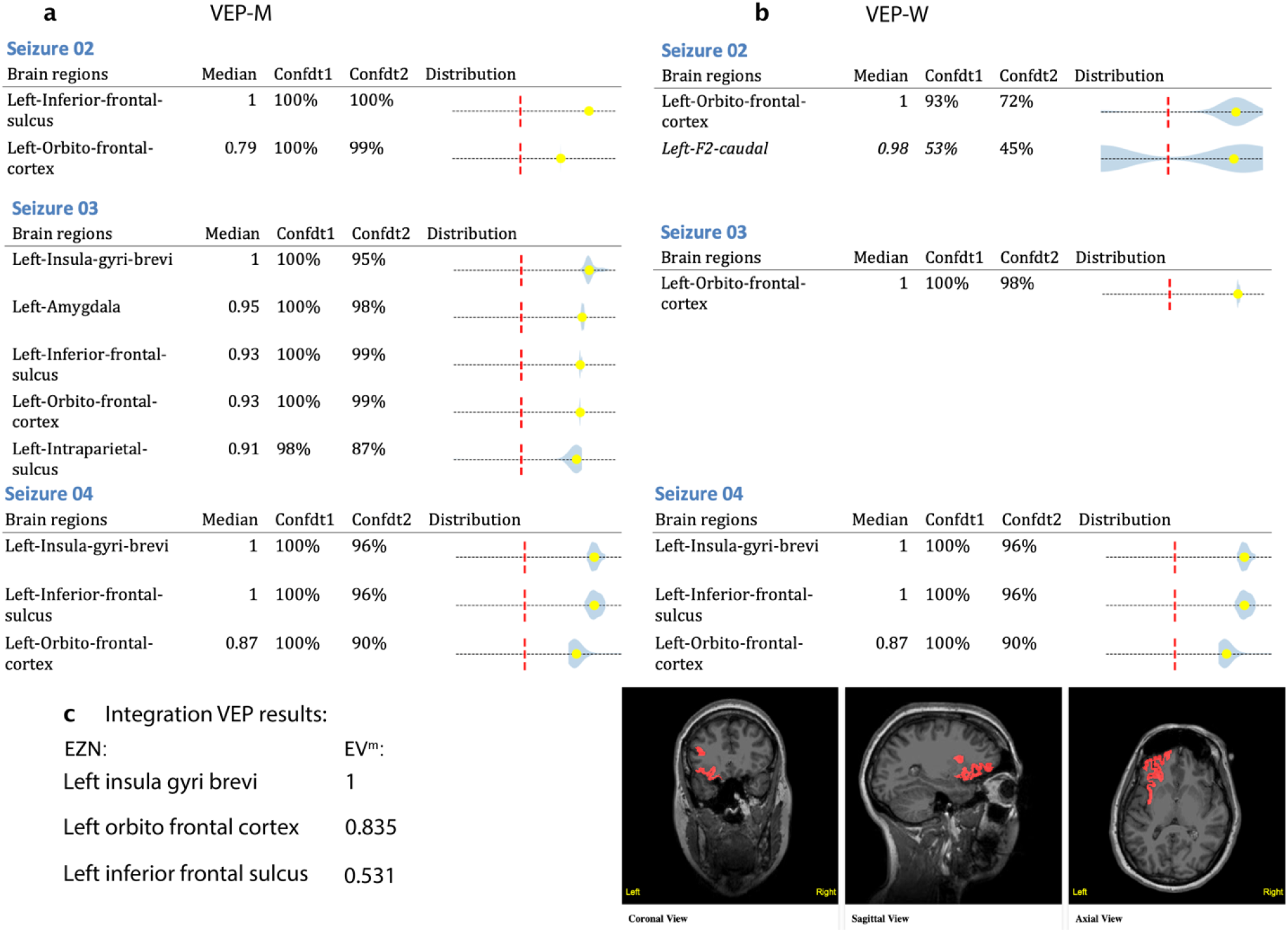
The clinical tables for the VEP results based on 3 more seizures for the example patient. a, With VEP-M prior. b, With VEP-W prior. c, Using the VEP results obtained by integrating 4 seizures from both VEP-M and VEP_W runs, we obtained the EZN for this patient: Left insula gyri brevi, left orbitofrontal cortex, and left inferior frontal sulcus. The numbers on the right are the integrated EV value. These three brain regions are highlighted in red in T1-MRI.

**Supplementary Fig. 10.**
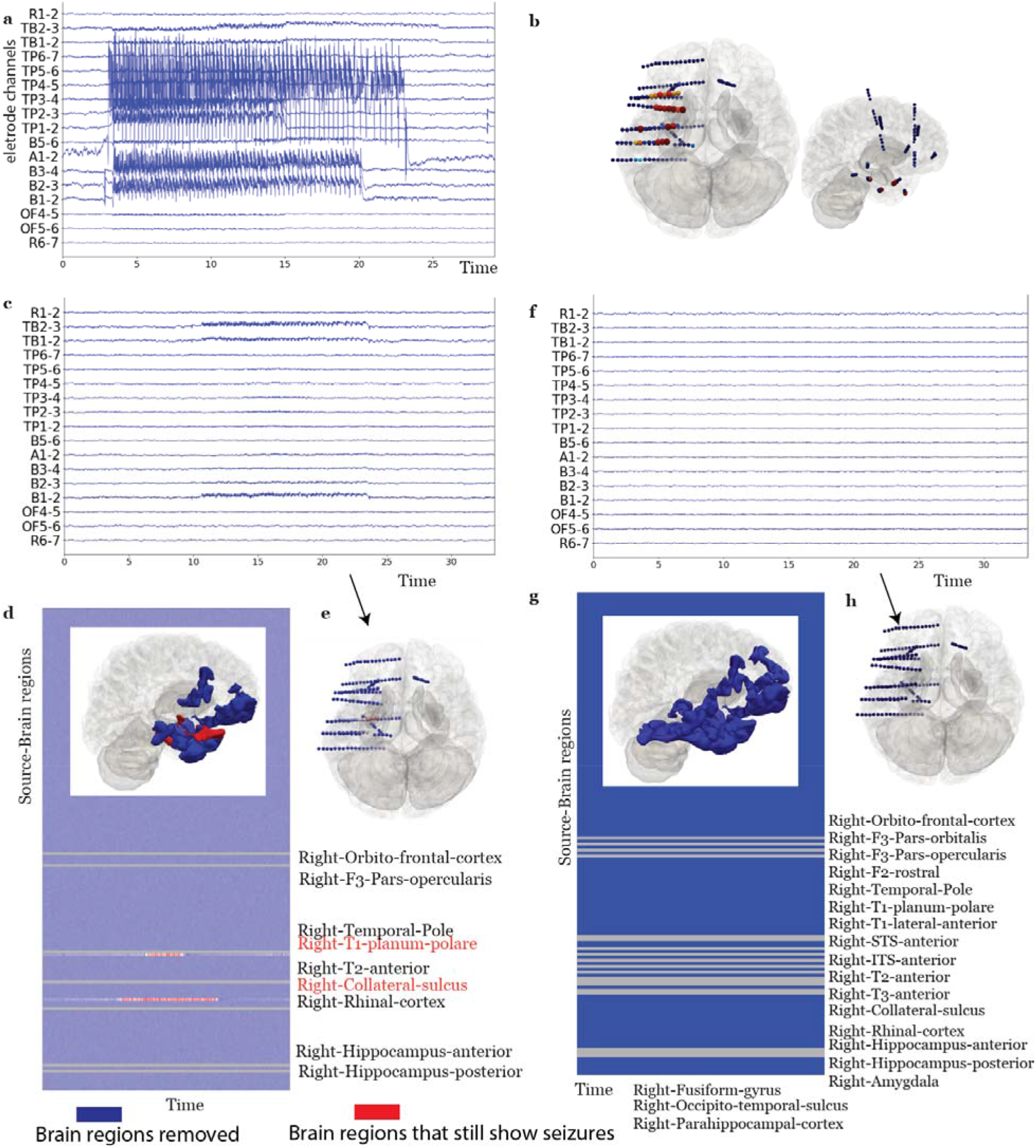
VEP virtual surgery for a patient with surgery seizure freedom. **a**, Selected simulated SEEG time series using the 6D-Epiletor with inferred parameters from the VEP results. **b**, Distribution of the signal power across contacts in a 3D brain from two different perspectives using simulated data as in **a. c**,**d**,**e**, Virtual surgery removing 7 regions in the clinical hypothesis. **c**, Simulated SEEG time series after virtual surgery. **d**, Time series and 3D brain in the source space; the 7 regions in blue are resected and listed in black; the red region indicates the remaining seizure activity. **e**, Distribution of the signal power across SEEG contacts of virtual surgery simulation in 3D brain. Same as **f, g, h**, but for the virtual surgery that entailed removing 19 regions in the real surgery.

**Supplementary Fig. 11.**
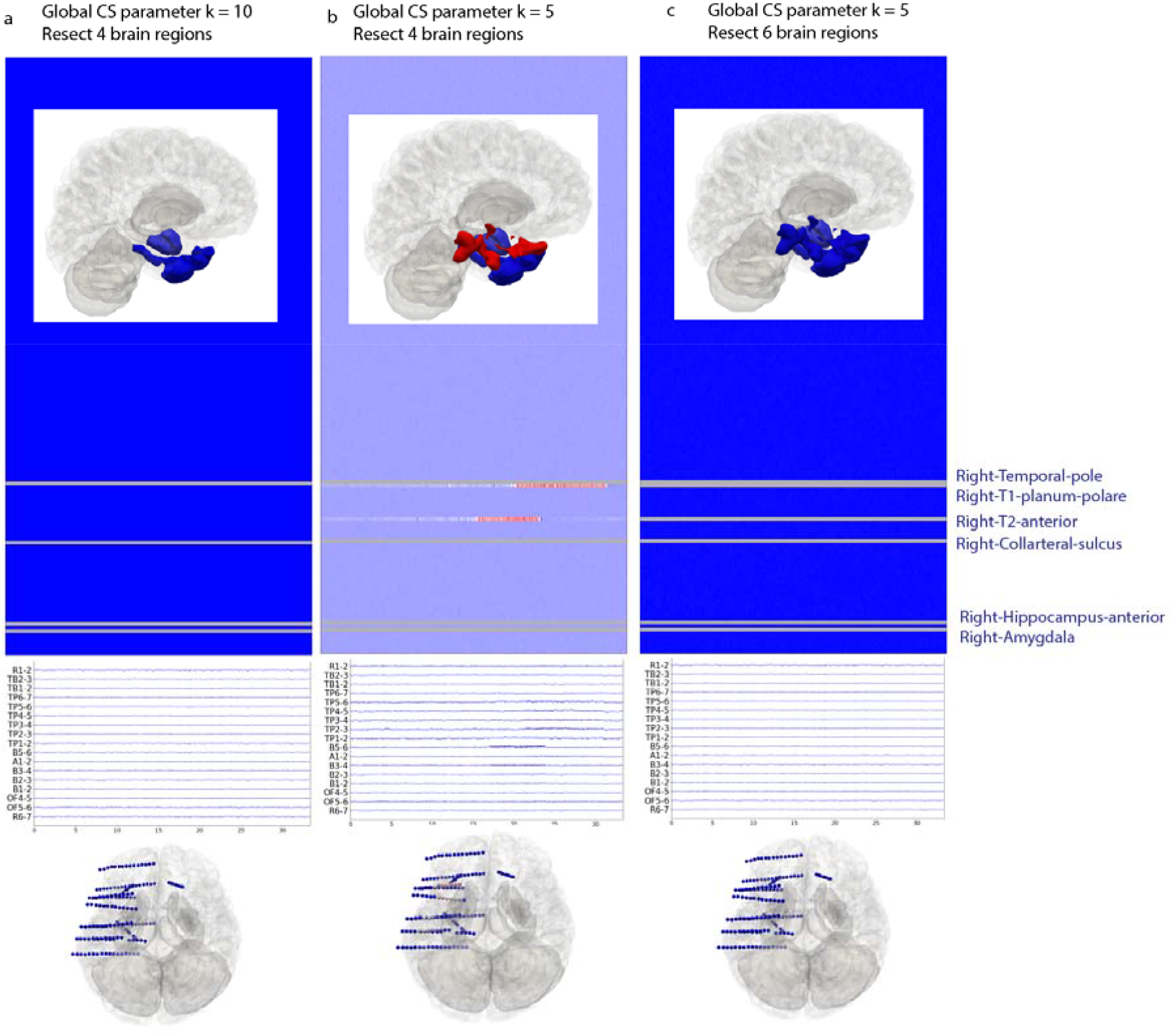
Three more virtual surgeries under two global connectivity scenarios for the same patient as Supplementary Fig. 9. **a**, If the global connectivity coupling parameter is *K* = 10 and 4 brain regions are resected, then the patient would achieve seizure freedom. The 4 resection regions are: right temporal pole, right collateral sulcus, right hippocampus anterior, and right amygdala. **b**, If the global connectivity coupling parameter is *K* = 5, the resection of the same four brain regions cannot allow the patient to become seizure free. **c**, If the global connectivity coupling parameter *K* = 5 and 6 brain regions (listed in the right) are resected, then the patient would achieve seizure freedom. For each sub-figure, the top is the time-series for the region source space and 3D brain with the resected regions highlighted in blue and the remaining seizure regions, if any, in red. The second panel is for the time-series in the electrode sensor space and the bottom panel is the power distribution among the electrodes.

